# Using a household structured branching process to analyse contact tracing in the SARS-CoV-2 pandemic

**DOI:** 10.1101/2021.02.03.21250992

**Authors:** Martyn Fyles, Elizabeth Fearon, Christopher Overton, University of Manchester COVID-19 Working Group, Tom Wingfield, Graham F Medley, Ian Hall, Lorenzo Pellis, Thomas House

**Affiliations:** Department of Mathematics, University of Manchester, UK; The Alan Turing Institute, London, UK; Centre for the Mathematical Modelling of Infectious Disease, London School of Hygiene and Tropical Medicine, UK; Department of Global Health and Development, London School of Hygiene and Tropical Medicine, UK; Department of Clinical Sciences and International Public Health, Liverpool School of Tropical Medicine, Liverpool, UK; Tropical and Infectious Disease Unit, Liverpool University Hospitals NHS Foundation Trust, Liverpool, UK; Department of Global Public Health, Karolinska Institutet, Stockholm, Sweden

## Abstract

We explore strategies of contact tracing, case isolation and quarantine of exposed contacts to control the SARS-CoV-2 epidemic using a branching process model with household structure. This structure reflects higher transmission risks among household members than among non-household members, and is also the level at which physical distancing policies have been applied. We explore implementation choices that make use of household structure, and investigate strategies including two-step tracing, backwards tracing, smartphone tracing and tracing upon symptom report rather than test results. The primary model outcome is the effect on the growth rate of the epidemic under contact tracing in combination with different levels of physical distancing, and we investigate epidemic extinction times to indicate the time period over which interventions must be sustained. We consider effects of non-uptake of isolation/quarantine, non-adherence, and declining recall of contacts over time. We find that compared to self-isolation of cases but no contact tracing, a household-based contact tracing strategy allows for some relaxation of physical distancing measures; however, it is unable to completely control the epidemic in the absence of other measures. Even assuming no imported cases and sustainment of moderate distancing, testing and tracing efforts, the time to bring the epidemic to extinction could be in the order of months to years.

## Background

The COVID-19 pandemic, arising from infection with SARS-CoV-2, has rapidly spread across the world leading to significant loss of life, with many different interventions employed in attempts to reduce the spread of the disease. To avoid overwhelming the capacity of healthcare systems and to limit mortality, many countries, including the UK, adopted policies to dramatically reduce the number of contacts via which infections could occur^2^. These ‘lockdown’ policies have included: requiring all but essential workers to stay physically in their residences; schools, universities, entertainment venues and all but essential businesses to close; and limitations on outside-household activities and meetings. In many countries, including England, there were dramatic curtailments in the number of outside-household social contacts made across most of the population but household contacts remained^2^. While effective in reducing epidemic growth, strict lockdown policies are not sustainable economically and socially^3^. As stricter regulations are relaxed, workplaces, schools and businesses reopen (albeit with additional safety measures), with increased contact between households occurring. In this context, the role of contact tracing and isolation is to target isolation to households with higher risk of infection to allow increased economic and social interaction without a return to rapid epidemic growth.

Contact tracing policies consist of three main components: *isolation* of identified infected individuals to prevent onwards transmission, *tracing* of their recent contacts who might have been exposed to infection, and the *intervention*, most commonly *quarantine*, applied to the traced individuals to try to halt the chains of transmission. As of August 2020 in the UK, a contact is defined as a case’s household member or a sexual partner and/or someone with whom they have: had skin-to-skin contact; coughed on; been within one metre of for more than one minute; had a face-to-face conversation with within one metre; been within two meters of for at least 15 minutes; or shared a vehicle with (or sat near if a plane or large vehicle). In the UK, case isolation and contact quarantine is done at home, even though individuals are advised to distance themselves from household members. Given the likely difficulties in achieving this in practice, contact tracing and isolation policies will likely not prevent within household transmission once an infection is introduced, but rather are aimed at reducing transmission between households.

Previous research on other respiratory infections finds households to be key structures in understanding population level transmission dynamics^4^. The nature and extent of contact between household members increases the likelihood of transmission compared to other types of social contacts. Households bring together disparate social networks such as those of different workplaces and schools, and they often bring together people of different age groups via family relationships. For SARS-CoV-2, emerging evidence is that household contacts of cases are much more likely to become infected than non-household contacts and a recent systematic review estimated household secondary attack rates to lie between 15.4-22.2%^5^. However, the reliance on symptomatic diagnosis of cases in many studies means this is likely to be an underestimate. Household structure should also benefit the contact tracing process in that household contacts are much more easily identified, so models or policies that do not account for this might miss these potential efficiencies. Given that negative serial intervals have been observed for SARS-CoV-2^6^, it is possible that a non-index case in a household epidemic develops symptoms before the index case, leading to a reduced time until symptoms are reported for the household branching process, compared to an individual level process where symptoms must be reported at each step.

Here, we use a household structured model to explore the potential impact of contact tracing and isolation on epidemic growth, considering a range of social contact, transmission, tracing performance and population adherence assumptions. We model the effects of potential strategies to improve contact tracing effectiveness, and consider the timeframe over which interventions must be held consistent in order to eliminate infection.

### Challenges to contact tracing and isolation

There are a number of challenges to using contact tracing and isolation to suppress SARS-CoV-2 transmission. Early models, even those made prior to upwards revision of unconstrained growth estimates^7^, suggested that contact tracing must operate with minimal delay and with high levels of accuracy (70-90% of contacts traced for R0 2.5-3.5)^8,9^ if it is to interrupt enough transmission chains to achieve control. The first step of identifying an index case is challenging for SARS-CoV-2; many cases are asymptomatic, and symptomatic cases frequently have mild or non-distinguishing initial symptoms. Once identified, the index case is isolated and subsequently interviewed to identify all individuals who may have been exposed while the index case was infectious, which becomes more difficult the longer the pre-symptomatic infectious period. Asymptomatic infections will not become index cases unless they get tested regardless of their symptom status. Some subpopulations, such as healthcare workers, might receive regular testing regardless of their symptom status, and asymptomatic cases may become index cases through this avenue, but this is the exception not the rule. The contact tracing interview with an identified case is subject to the delay between infection and symptom onset and/or case confirmation via testing, plus any further procedural delay to the interview. The task of reaching the contacts is also subject to delays and potentially impossible when they are non-identifiable (e.g. strangers to the case), unreported (e.g. if the case fears disclosure of their contacts or the contact tracing process is not adapted to the needs of the population^10^) or unreachable. For a new infection, it is also possible that the contact definition used for tracing is not well matched to the main modes of transmission. Traced contacts are then asked to quarantine to restrict the potential spread of the infection, and are potentially monitored for signs of infection. If a quarantined contact meets the case definition or develops symptoms, their contacts prior to quarantine are in turn then the subjects of tracing attempts.

Timing of symptom onset versus infectiousness is important^11,12^. Control of SARS-CoV-1 was facilitated by the fact that peak infectiousness occurred after the onset of noticeable symptoms and that there was little pre-symptomatic or asymptomatic transmission^13,14^. For SARS-CoV-2, significant pre-symptomatic transmission means that by the time an infected individual is identified, there is a high probability that they have already infected others^15–17^. Many infected people are asymptomatic or experience mild symptoms, remaining undiscovered^18^. This issue is compounded by limited testing and high rates of false negative in tests to diagnose active infections^19^.

Identified contacts in the UK are asked to quarantine for a period of 14 days following contact with the case (within which the vast majority of incubation periods would occur^20^). If the contact develops symptoms and tests positive, they must self-isolate for 10 days and their contacts, including those in their households, must quarantine for 14 days from the time of their symptom onset^21^. Particularly in the absence of contact testing and release from quarantine if negative, there will be a proportion of society that is unable or unwilling to take up or adhere to issued notices to quarantine or to isolate for the full period. The effects of non-uptake or of partial adherence on epidemic growth will depend on the dynamics of within-household transmission, making this important to capture in modelling. There might also be practical trade-offs since a strict system that in theory might be most effective in reducing transmission could result in lower adherence; and there are complications to releasing contacts who test negative early due to low test sensitivity^22^. While there is evidence that the average number of contacts made at the beginning of the UK’s ‘lockdown’ at the end of March 2020 was reduced by over 70%^23^ compared to a 2006 contact survey^14^, another survey from two days in early May 2020 found that almost three quarters of individuals who showed or whose household showed SARS-CoV-2 symptoms had left the house in the previous 24 hours, behaviour that was contrary to policy at the time^24^. Across past studies, reporting of adherence to quarantine orders varies widely, and generally involves shorter time frames than those involved in the current pandemic. Further, we would expect adherence to vary over time as the perceived risk of breaking quarantine, public trust in health services and government, and the benefits of breaking quarantine (such as socialising, which may be impossible under lockdown) change over time. In practical terms, this means that we will not know what adherence levels might be expected over the time period necessary for control of SARS-CoV-2 in the absence of a vaccine^25^.

### Contact tracing, isolation strategies and considerations post-‘lockdown’

Those countries that have maintained periods of SARS-CoV-2 epidemic suppression have used contact tracing, isolation and quarantine measures alongside a variety of other interventions^26,27^, and have re-deployed stricter physical distancing again in the event of rising cases. This experience is consistent with modelling that suggests that such suppression requires contact tracing to be implemented alongside policies that reduce usual patterns of social contact^28^.

There are modifications to the contact tracing process that could improve efficiency and effectiveness. The strong likelihood of pre-symptomatic transmission within the household means that tracing not only the index case’s contacts but contacts of the whole household, and quarantining the whole household of a contact might be effective. Immediate quarantine of contacts rather than monitoring for symptom development contributes to prevention of asymptomatic transmission and pre-symptomatic transmission if tracing occurs quickly enough^29^. Immediate tracing and quarantining of contacts-of-contacts - ‘two-step tracing’, implemented for some contacts as part of Vietnam’s strategy^26^ - could further improve the speed of the control process relative to transmission, but at the cost of a large number of people being quarantined per case, which if widely applied may lead to effectively placing small areas in lockdown^30^. In the long run, this initial cost might be worthwhile if better epidemic control is achieved. Delays due to testing can be eliminated if tracing is initiated upon symptom reporting, though in practice many with symptoms will not actually have SARS-CoV-2 and a sizable proportion with SARS-CoV-2 will have no symptoms^31^. To reduce tracing delays and increase the number of contacts who can be identified and traced, the use of smartphone apps that record proximate devices (a proxy for contacts) and can send them notifications to quarantine upon a case testing positive, have been explored in a number of settings and trialled on the isle of Wight^32^. Modelling suggests that such an app could potentially bring the effective reproduction number *R*_eff_ below one, but only if population uptake is very high^9^. In the UK, this would equate to at least 80% uptake by smartphone owners^10^, which is higher than that yet achieved elsewhere. Fears over privacy, stigma and misinformation could impede uptake^33^. In countries which have deployed contact tracing applications the uptake is very mixed; data on app uptake was collected from news articles for 30 countries, with 19/30 countries achieving uptake of less than 10%^34^.

There is an implicit form of direction in contact tracing - an individual is said to be *forward* contact traced if the direction of contact tracing is the same as the direction of transmission, which results in the traced individual being an infectee of the case from whom the tracing attempt was initiated. *Backwards* contact tracing is when the direction of tracing is opposite to the direction of transmission, which results in the traced individual being the infector of the case who initiated the tracing attempt. For some diseases, backwards tracing can be an important goal as once you have traced backwards you can then trace forwards again to discover ‘sibling’ infections who share the same infector^35^. A visualisation of backwards tracing is plotted in Figure 1, where a successful backwards tracing event leads to the quarantine of a sibling chain of transmission.

**Figure 1:**
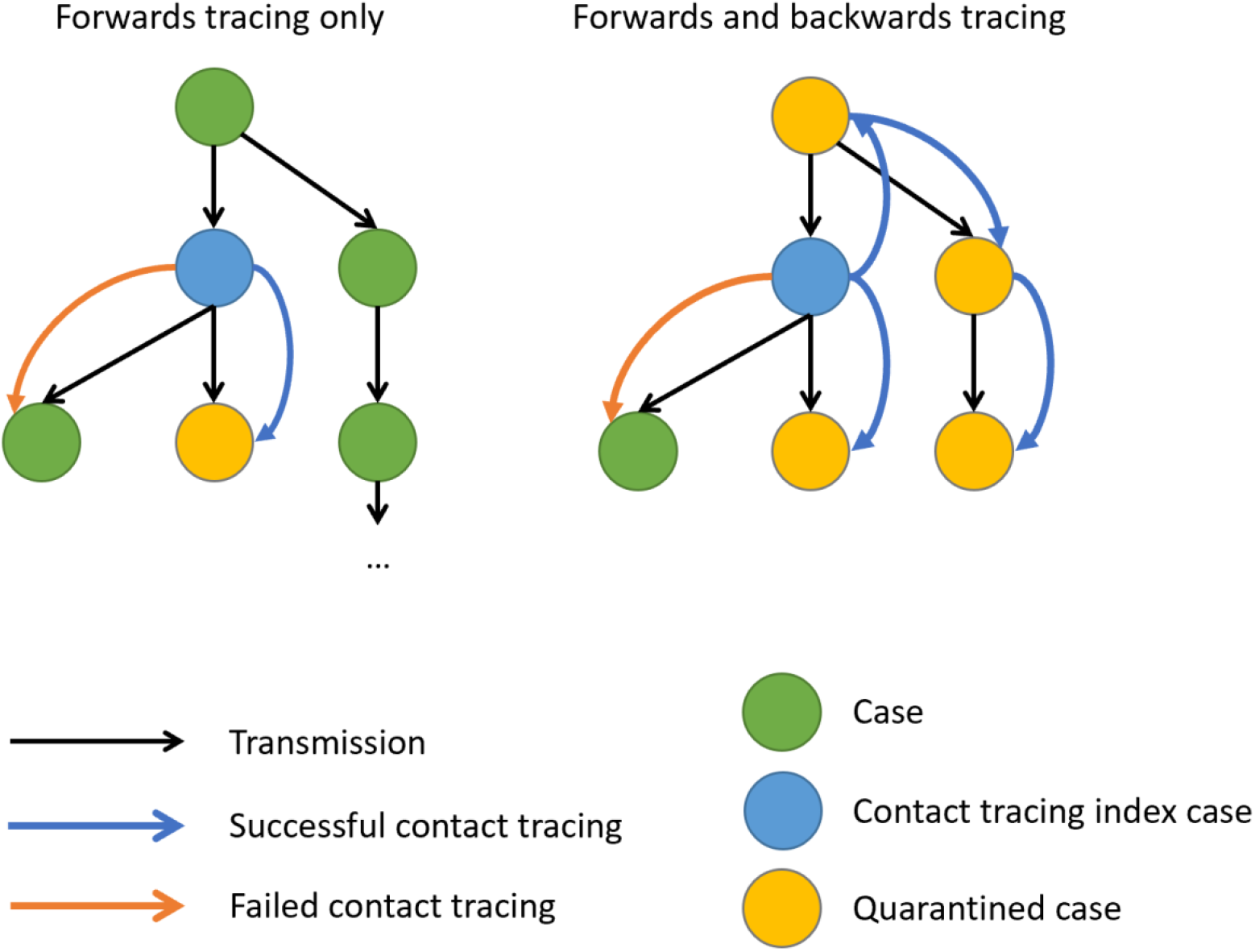
A demonstration of backwards and forwards contact tracing. Here, the chain of transmission is represented using black arrows. The blue circle represents the first case in this chain to be detected. The left hand contact tracing chain contains only forwards contact tracing and as a result only the infectees of the index case are traced. The right hand plot has the same forwards tracing as before, and a backwards contact tracing event in which the infector of the index case was discovered, and forwards contact tracing then enables sibling infections to the index case to be traced and quarantined.

Implementation of backwards tracing in practice requires increasing the time window back from symptom onset over which contacts are traced to include the likely date of the case’s infection, not only onset of their infectiousness. Backwards tracing might be especially efficacious due to a biased sampling effect; the more secondary infections an individual causes, the more likely they are to be isolated through backwards tracing initiated by one of their secondary infections. Therefore backwards tracing has an increased likelihood of finding individuals who caused a lot of secondary infections, and for these individuals it is then highly worthwhile *forwards* tracing to find their infectees^36^. Following this logic, if the number of secondary cases is highly overdispersed, as has been estimated^37^, then backwards tracing could be especially useful to help identify ‘superspreaders’ and transmission chains that they initiate^35^. However, tracing backwards to move forwards again might be less useful when tracing delays are high relative to the speed of transmission^38^, and when accurate recall or identification of contacts degrades over longer time frames.

If it is decided that the goal of contact tracing is to achieve elimination, then to inform decision-making, we need to understand the time scales over which contact tracing and concurrent physical distancing interventions must be sustained to reach elimination, without which transmission is likely to climb again. Even if interventions are enough to drive the infection to elimination, we might expect a long ‘tail’ of ongoing but increasingly sporadic transmission during which time reintroduction of the epidemic is an ongoing risk if tracing and physical distancing measures are relaxed^39^.

Here we consider what role contact tracing and isolation might play in suppressing epidemic growth across a range of plausible epidemiological and behavioural assumptions. We begin more generally and then consider the case of the UK more specifically during May-June 2020, considering lockdown easing scenarios and the contact tracing and isolation policy adopted on May 28th 2020. To capture the interplay of within- and between-household epidemic dynamics^40^ we use a household-structured branching process model to investigate how contact tracing and isolating might affect the growth rate, the probability of epidemic extinction and extinction times for: household versus individual-based tracing strategies; different levels of physical distancing; assumptions about the probability of discovering infected individuals; a range of suggested improvements to contact tracing, including an app to reduce tracing times and improve the probability of identifying and reaching contacts. We also examine how key aspects of implementing the interventions, as well as uptake and adherence to them among the population, might affect their efficacy in practice.

## Methods

### Model

#### Household-Individual Branching Processes and Contact Tracing

We model the SARS-CoV-2 epidemic as a branching process of infections of individuals grouped into households. Transmission is considered on two levels: *local* within-household transmission and *global* between-household transmission. A static population of households, excluding migration in and out of households, is assumed and infection can be introduced into a household only once. In mathematical terms, a household-structured branching process is considered to be a multitype branching process, where the type of an individual is the generation of the household epidemic they belong to in combination with their household size. The number of offspring produced by a case is conditional upon the size of the household they belong to, and how many susceptibles remain in that household. Upon introduction of the infection to a household, a within-household epidemic is initiated, referred to as the *local epidemic*.

Individuals infected by a local epidemic are able to propagate the infection globally by introducing the infection to a fully susceptible household. Because the virus is always introduced to a fully susceptible household, the model represents a worst case scenario of no immunity. Individuals in the model differ only by the size of the household they belong to, and we do not model explicit household compositions (e.g. by age, gender). Seroprevalence in the UK in May-June 2020, was estimated to range from 0.5% to 17.5% in the worst affected region, with immunity low overall in the UK.^41^.

The transmission tree generated by the branching process allows for modelling of contact tracing as a type of ‘superinfection’ along the tree; for a household-individual branching process the contact tracing process superinfects at the level of households (Figure 2). When the contact tracing process ‘superinfects’ a household or individual, then the effects of the contact tracing process are applied. These are typically: quarantine, resulting in a reduced ability of cases in the household to transmit the infection globally; and surveillance, where all individuals in the household are on the lookout for symptom onset.

**Figure 2:**
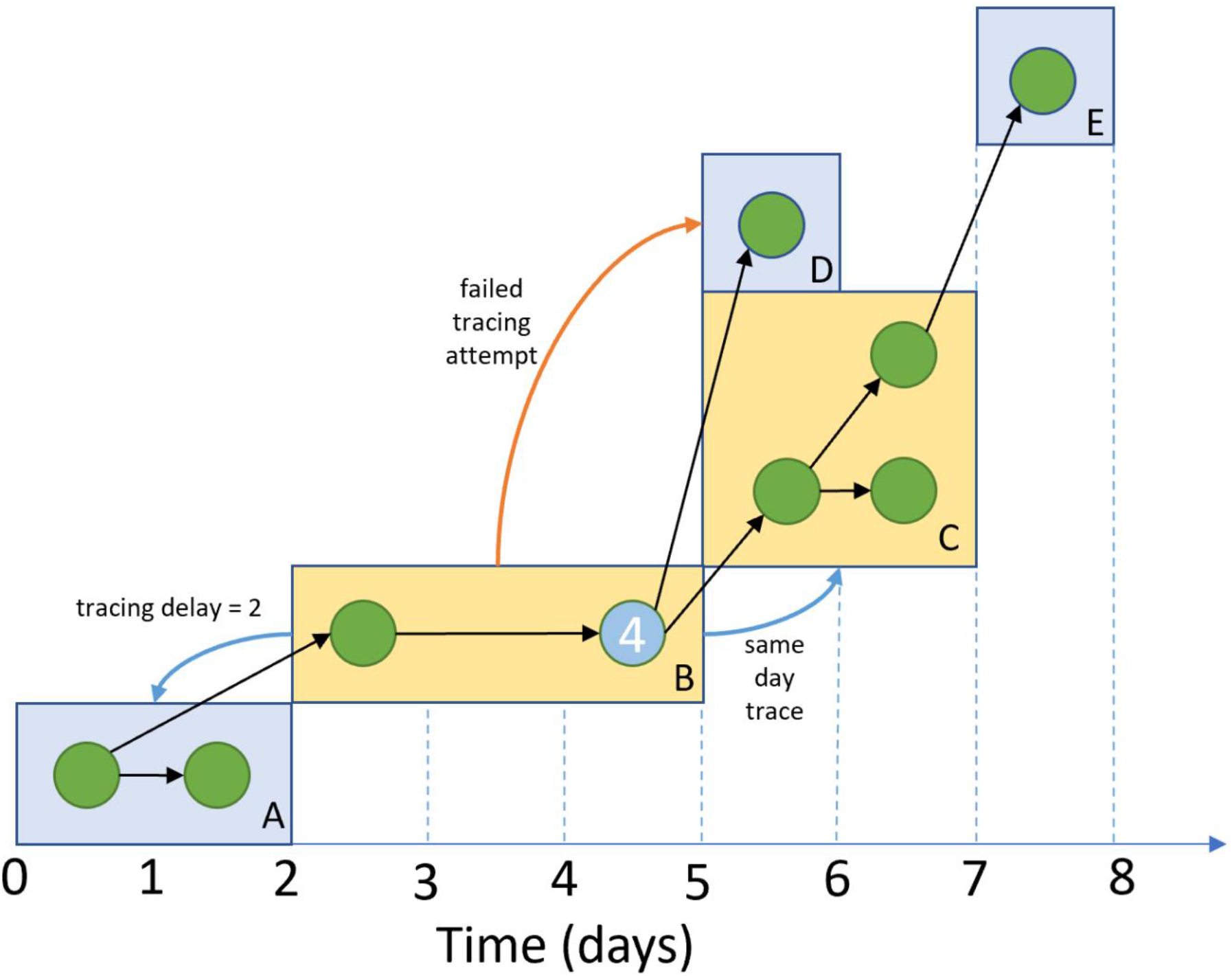
Example of the transmission and household tracing and quarantine process. A demonstration of a household branching process with a contact tracing process, the households are identified by letters in the bottom right hand corner of each rectangle. The infection is discovered in case 4. This quarantines household B and initiates contact tracing of connected households A, C and D. The backwards tracing attempt to household A succeeds, with a time delay of 2 days. Household C is traced immediately, quarantining several cases early in their infection. When there is symptom onset in one of these cases the contact tracing process will propagate again by attempting to reach household E, potentially after a testing delay. Household D was not successfully traced by household B, and will continue to spread the infection. The x-axis refers to the temporal evolution of the transmission process in this example.

The household structure introduces additional effects to the contact tracing dynamics that enable the exploration of household as well as individual-level contact tracing and isolation policies. Each infection in a household has an independent and identically distributed probability of being identified and reported, and therefore, infection is more likely to be identified in larger households. Larger households tend to make more global contacts than smaller ones, and therefore may be responsible for spreading more of the infection, but are also more likely to have an infection identified in them. Later generations of a local epidemic may be quarantined early in their infection due to a household member belonging to an earlier generation of the local epidemic identifying and reporting the infection.

At time 0, the infection is seeded by creating a number of initial infections with infectious age 0, as to do otherwise would change the dynamics of the local within household epidemics for the first generation households.

The household branching process approximation is derived when the epidemic is considered to be spreading through a population of individuals segmented into households, with homogenous mixing between households. If it is early in the epidemic, and there is little to no susceptible depletion then the homogenous mixing assumption implies that the probability the virus is reintroduced into a previously infected household is effectively zero. The model is therefore limited in that the approximation does not hold when there is significant susceptible depletion, and for this paper we therefore take the population size to be infinite. The branches refer to the fact that we ‘connect’ two cases if there was a transmission between them, and this forms chains of transmission.

#### Social Contacts and Infection

Each day, individuals make contacts, the number of which is sampled from an overdispersed negative binomial distribution (overdispersion parameter 0.32; see Supplementary Material for technical details) estimated with data on ‘all contacts’ (i.e. physical and non-physical) from the 2006 POLYMOD study^42^, stratified according to the size of the household in which the individual resides. Household sizes are aligned with the 2019 ONS survey^43^, and are categorised as of size 1, 2, 3, 4, 5, 6+. Contacts are only distinguished by whether they are within (local) or outside (global) the household, with the proportion of each again estimated using POLYMOD. A local contact with a household member occurs each day with fixed probability, conditional upon household size. As such the number of local contacts is binomially distributed. The number of global contacts is given by the difference between the total contacts made and the local contacts. If an illegal combination of contacts is observed (e.g. three local and two total) then all contacts are redrawn from their distributions until a legal combination occurs. We do not consider repeated global contacts, so individuals experience no susceptible depletion among those whom they might infect globally. We do not consider the extent to which each household member’s global contacts overlap. Again, this represents a worst case scenario for epidemic spread, although not necessarily for the efficacy of contact tracing^44^.

Physical distancing and lockdown measures are modelled through a Bernoulli thinning of an individual’s global contacts, with their local contact remaining unchanged, leading to a percentage reduction in global contacts made. For example, suppose that physical distancing leads to a 50% reduction in global contacts, then we would draw the number of global contacts that an individual would make in the absence of physical distancing, and then consider each contact to occur with probability p=0.5. This leads to a binomial number of actual contacts made conditional upon the number of trials being drawn from an overdispersed negative binomial, (see supplementary information for details). We explore physical distancing by applying the same reduction in global contacts to every individual, which we refer to as uniform physical distancing. Because we expect there to be some dependence upon household size and composition in practice, we later assume lockdown relaxation scenarios and derive physical distancing parameters for these scenarios where the physical distancing reduction is conditional upon the size of the household.

Because the probability of a contact causing infection cannot be directly observed, we condition the probability that a contact spreads the infection upon the infectious age of the individual, in such a way that generation times are Weibull-distributed with a mean of 5 days and standard deviation of 1.9 ^9^, where a generation time is defined as the delay from becoming infected to a transmission of the virus. The distribution of generation times is an important quantity to calibrate, as it defines a variety of timings and quantities within the model, for example the infectious ages and quantity of secondary infections by the time that an individual tests positive. The probability of a contact leading to infection is tuned for the local and global settings such that the model has a baseline growth rate of *r* = 0.22 per day^45^ and a household secondary attack rate of 21.4%, based on the mean of several studies that were available during early April 2020^46–48^. The baseline scenario assumes that individuals activate self-isolation and household quarantine when they believe they are infected, but there is no active contact tracing process. Thus, any observed reduction in the growth rate can be attributed solely to the contact tracing process, and not to effects of the case’s household quarantine.

#### Case Identification and Contact Tracing Delays

For contact tracing to be initiated, an infected individual first develops symptoms over a gamma distributed incubation period with mean of 4.84 days and standard deviation of 2.78^20^. As an indicator of where the case discovery probability might lie, we consider the asymptomatic case probability, estimates of which range between 30-70% of cases^31,49–51^. We consider an upper limit in the case discovery probability of 50%, to account for mild cases that do experience symptoms but do not report them.

Case identification leads to an immediate quarantine of all other household members, and we explore initiating tracing both immediately upon symptom report and waiting for a positive test result before initiating tracing. We assume that once symptoms develop there is a delay before the case reports the symptoms, which only applies if the household is not quarantined. Quarantined households are aware they have been exposed and therefore report infection at symptom onset. If tracing begins after a positive test, we add a further gamma distributed testing delay of mean of 1.54 days and standard deviation of 1.1, parameterised using a truncation corrected maximum likelihood estimator applied to anonymised UK line-list data^20,52^.

Contacts are traced with a probability of success, and successfully traced individuals are assigned a tracing delay time from a Poisson distribution, with a mean value that we vary between 1.5 and 2.5 days. We explore a range of possible contact tracing success probabilities from 70-95%, the upper bound of which was the percentage of known contacts (i.e. those who could be identified) who were successfully traced by Public Health England during the containment phase^53^. At the end of the tracing delay, the successfully traced household is quarantined. If an individual is adhering to quarantine or isolation, we assume they make no global contacts at all, but within-household contacts continue, though do not occur more frequently when outside-household contacts reduce. Therefore, we assume no increase in local transmission while a household is quarantined.

#### Contact Tracing Strategies

Our default contact tracing strategy is ‘household-based’ and designed to take advantage of the household structure; when an infection is identified in a household all members of the household have their contacts traced. The rationale behind household level contact tracing is that once a case has been detected in a household, there is an increased likelihood that their household members are already infected and have possibly spread the infection outside the household, but have not had symptom onset or received a test result. Thus, household level contact tracing would initiate contact tracing earlier in the infection for cases who were infected by a household member. When a contact is successfully traced, their whole household quarantines. We also compare this to an ‘individual-based’ contact tracing process in which only individual cases have their contact traced, and only the specific people identified as contacts and successfully traced are quarantined. The individual-based model, with household structure affecting infection dynamics, but quarantining of contacts implemented on an individual level, is closest to what was adopted in the UK in May 2020.

Given the speed of transmission, we further consider a ‘two-step’ contact tracing process at the level of households, as illustrated in Figure 3. One step contact tracing attempts to isolate individuals who are distance 1 from a known case, and two-step contact tracing attempts to isolate individuals who are distance 2 from a known case. As previously discussed, contact tracing can either occur at the household level or the individual level, and this leads to slightly different distances as the fundamental epidemiological unit is different. For household level contact tracing, individuals are distance 1 if they live in different households, but there has been contact between the households by any two members. Variations on this approach have been used in Vietnam for SARS-CoV-2^26^, although typically only on the level of individuals.

**Figure 3:**
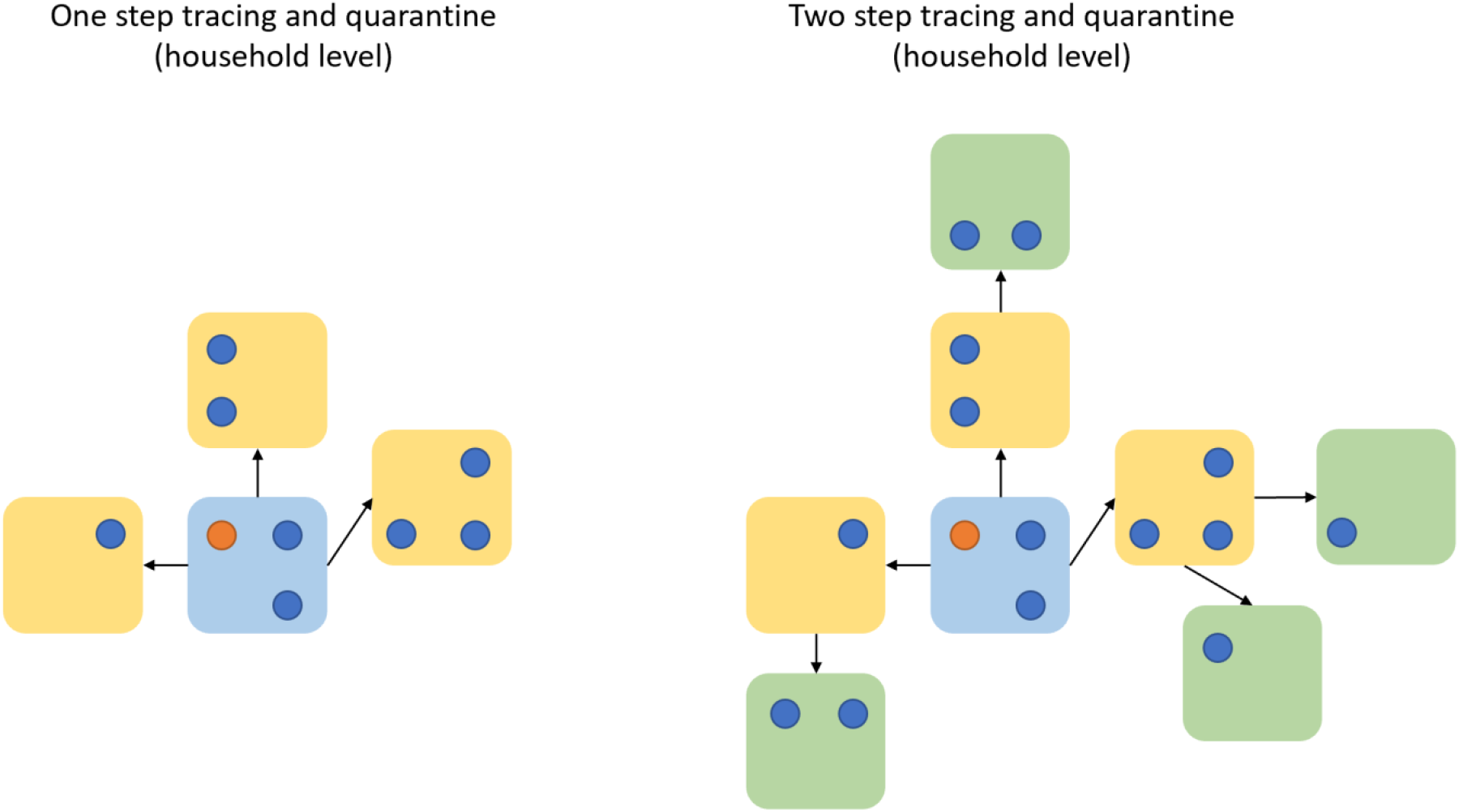
A demonstration of two step tracing on households. We assume that all within-household contacts are always traced. For both one and two step tracing at the household level, all household members undergo quarantine and tracing regardless of whether they are the index case or the contact of the index case. In both of these examples, all of the individuals have been placed under quarantine or isolation once detected (for the index case) or traced. Households who were not successfully traced are not shown here.

To consider a smartphone tracing app scenario^54^, we allocate a random proportion of the population to be running the app on their smartphones, considering a wide range (Table 2). When both individuals on the ends of a transmission event have been allocated the app, the transmission is traced with 100% probability, and the tracing delay is reduced to 0. As both ends of a transmission event are required to have the application, the probability of seeing the app trace the transmission event is quadratic in the probability of an individual having the app. Here, we are considering a ‘perfect world’ contact tracing app, and making the assumption that the app is able to record all epidemiologically relevant contacts. We do not allow for the possibility that the app misses a contact which causes a transmission.

In our models exploring the effect of adherence to quarantine, we consider 1) non-uptake of quarantine among traced contacts, implemented at the level of households; and 2) declining adherence to quarantine over time, whereby a proportion of households have a propensity for declining adherence^24^. Among these households, each individual has a fixed probability of leaving quarantine each day, and can then make global contacts/infections again.

A practical implementation of backwards tracing is considered, as it relates to the strategy implemented in the UK. The probability of including the infecting individual on a list of contacts to be traced depends on how many days pre-symptom onset are included as days when eligible contacts were made. For instance the UK policy traces back 2 days in recognition of the significant role that pre-symptomatic transmission plays, but this is unlikely to include the infecting individual, given the distribution of incubation periods (with an approximately 5 day mean). To include the infecting individual as a potentially traceable contact, this time window would need to be up to 14 days. However, the further ago a contact was, the more difficult it might be to recall. A simple model of recall is applied, where the ability to recall a contact decays at a geometric rate as a function of the number of days since the contact occurred. There is little evidence to inform this parameter; our assumed parameters come from the personal experience of contact tracers. If the contact tracing is performed digitally through the app, then there is no recall decay and we explore the interaction between digital contact tracing and recall decay.

### Parameters

Parameter values used for our simulation and their sources are summarised in Tables 1 and 2. We distinguish between several types of parameters; data driven, uncertain and scenario parameters. Data driven parameters (Table 1), such as the household size distribution, are not included in the sensitivity analysis distribution as they cannot be varied in a sensible way. Uncertain parameters such as the probability of contact tracing success are subject to significant uncertainty and we sample these from a prior distribution between simulation runs to perform a sensitivity analysis. In several sections of this paper, we choose parameters that represent fixed scenarios that are chosen to make the results more interpretable. For example, when we examine increasing the amount of backwards tracing that is performed then we keep the epidemic and contact tracing parameters fixed. We assume fixed parameters when we are investigating backwards tracing are detailed in Table 3. Our assumed scenarios of lockdown relaxations are detailed in Table 4, and the resulting reduction in social contacts stratified by household size are described in Table 5.

**Table 1:** Data driven parameters.

| Parameter | Values | Source |
| --- | --- | --- |
| Baseline Growth Rate (pre-interventions) | 0.22 per day<br>(doubling time approximately 3 days) | 45 |
| Incubation period | Gamma (mean= 4.84 days, sd= 2.78 days) | 20 |
| Generation time | Weibull (mean=5, sd=1.9 days) | 9 |
| Household Size Distribution | 1: 0.29, 2: 0.35, 3: 0.15, 4: 0.14, 5: 0.05, 6+: 0.02 | 43 |
| Onset to isolation | Gamma(mean = 2.62 days, sd = 2.38 days) | Assumed using data from Singapore onset to visit to medical provider<sup>45</sup> |
| Recall decay rate | 10% | Assumed from experience of contact tracers |

**Table 2:** Sensitivity analysis parameters.

| Parameter | Value | Source | Sensitivity analysis distribution |
| --- | --- | --- | --- |
| Reduction in global contacts per day due to physical distancing | 0-90% | Lockdown reduction around 90%<sup>23</sup> | Uniform(0.0, 0.9) |
| Probability that an infected individual self identifies | 0.1, 0.2, 0.3, 0.4, 0.5 | Bounded using asymptomatic infection probabilities | Equal probability |
| Testing delay (isolation to confirmed positive) | Specimen to report delay, Gamma (mean = 1.5, sd = 1.11 days)<br>(only applies to simulations that require testing before tracing) | Estimates informed by NHS Test and Trace statistics<sup>55</sup> | Mean varied between 1.5 days and 2.5 days |
| Tracing delay | We model as a Poisson with mean distributed between 1.5 and 2.5 days | Estimates informed by NHS Test and Trace statistics<sup>55</sup> | Poisson Parameter ~ Uniform(1.5, 2.5) |
| Probability of contact tracing success for global | 70 - 95%<br>Based on 95.2% of all *identified* contacts successfully | PHE containment period contact tracing<sup>53</sup> | Uniform(70, 95) |
| contacts (in the absence of an app) | traced during UK containment period. |  |  |
| Proportion of population with a smartphone and with app installed | 0% to 50% | Singapore's Trace Together uptake of approx 40% (Sept 2020) <sup>7</sup>. Other apps reporting lower uptake<sup>34</sup>. | Uniform(0, 0.5) |
| Uptake of quarantine among traced households | 50%-100%<br>(only applies to simulations where non-adherence is allowed) | Assumed | Uniform (0.5,1) |
| Proportion of households that have the propensity to not adhere to full quarantine stay | 0% to 50%<br>(only applies to simulations where non-adherence is allowed) | Assumed | Uniform(0, 0.5) |
| Daily probability to leave isolation early (if household has the propensity to not adhere to full quarantine stay) | 0% to 5%<br>(only applies to simulations where non-adherence is allowed) | Assumed | Uniform(0, 0.05) |

**Table 3:** Model parameters for backwards tracing simulations.

| Parameter | Value |
| --- | --- |
| Infection reporting probability | 30% |
| Contact tracing success probability | 80% |
| Mean contact tracing delay | 2 days |
| Reduction in global contacts | 30% |
| Mean testing delay | 1.5 days |

We only vary the number of days prior to symptom onset are traced.

**Table 4:** Assumed lockdown relaxation scenarios.

| Scenario | Workplace Contacts | School Contacts | Leisure Contacts |
| --- | --- | --- | --- |
| A | 20% increase | 10% resume | 0% resume |
| B | 30% increase | 25% resume | 10% resume |
| C | 30% increase | 50% resume | 10% resume |
| D | 40% increase | 60% resume | 30% resume |
| E | 50% increase | 100% resume | 75% resume |

*The baseline number of cases is calibrated to the England lockdown of March-April 2020, and we consider increasing the number of contacts that occurred. We consider school and leisure contacts to resume to the pre-lockdown levels and consider small step increases in the number of work contacts.*

**Table 5:**
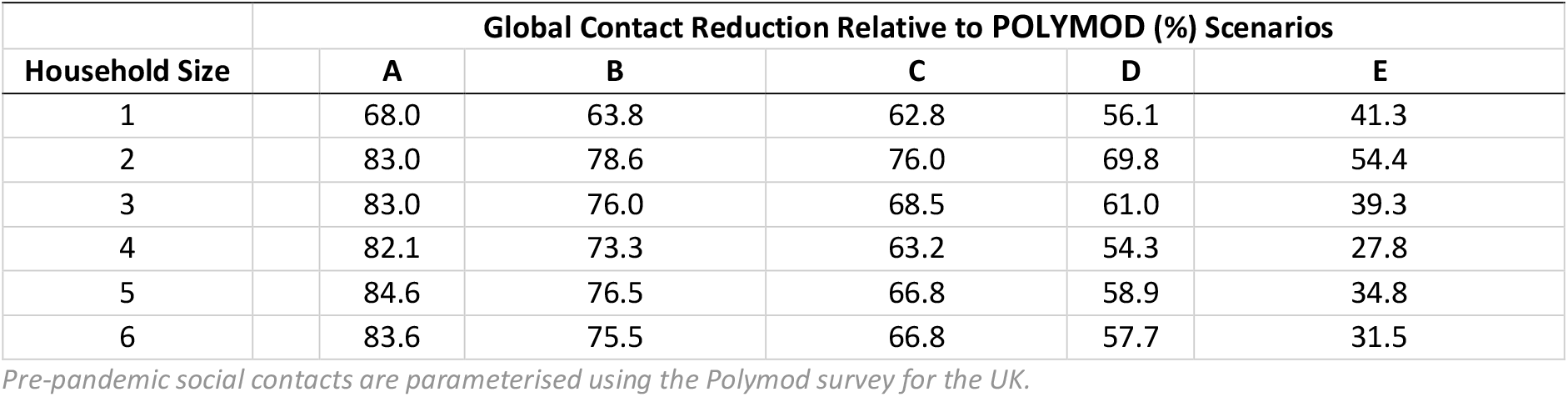
Global contact reductions relative to pre-pandemic social contacts by scenarios described in Table 4, stratified by household size.

|  | Global Contact Reduction Relative to POLYMOD (%) Scenarios |  |  |  |  |  |
| --- | --- | --- | --- | --- | --- | --- |
| Household Size |  | A | B | C | D | E |
| 1 |  | 68.0 | 63.8 | 62.8 | 56.1 | 41.3 |
| 2 |  | 83.0 | 78.6 | 76.0 | 69.8 | 54.4 |
| 3 |  | 83.0 | 76.0 | 68.5 | 61.0 | 39.3 |
| 4 |  | 82.1 | 73.3 | 63.2 | 54.3 | 27.8 |
| 5 |  | 84.6 | 76.5 | 66.8 | 58.9 | 34.8 |
| 6 |  | 83.6 | 75.5 | 66.8 | 57.7 | 31.5 |

*Pre-pandemic social contacts are parameterised using the Polymod survey for the UK.*

### Simulated Scenarios

#### Effects of household structure and contact tracing strategies and parameters on growth rates

To examine the effects of households on transmission and tracing dynamics, we first compare the relationship between growth rates and global contact reductions in models with and without household structure. We implement models with no household structure by giving each household in the model a size of one. For models with households, we compare the effectiveness of ‘individual-based tracing’ with ‘household-based tracing’, as described previously.

We estimate growth rate of epidemics by simulating 5000 infected cases to start on Day 0 with infectious age 0, which ensures there are enough infections to remove the probability that the epidemic immediately goes extinct and for improved estimation of the growth rate. The epidemic is simulated for 25 days and log-linear regression is used to estimate the growth rate during days 15 to 25. The first 15 days are discarded as model burn-in, so that the infectious ages of cases can become well mixed and for the contact tracing to have been initiated. We first compare the growth rate from a baseline scenario where symptomatic cases initiate self-isolation for themselves and their household members quarantine as described above, but there is no contact tracing initiated upon report of symptoms. Unconstrained growth rates in the absence of isolation or quarantine for different probabilities of case discovery against global contact reductions are shown in the supplementary materials.

Using a model with household structure and household-based tracing, we examine manual tracing, the effects of a hypothetical tracing app, and 2-step tracing across simulations with some parameters set as described in Table 1 and other parameters varied as described in Table 2. We explore the effects of some households not taking up quarantine or reducing their adherence over time, and we consider initiating tracing on test results rather than on symptoms. The resulting observations of the growth rate are variable due to the stochasticity of the simulations and the variability in the parameters, which are sampled from prior distributions as described in Table 2.

#### Simulation of extinction times and probabilities under contact tracing

Using the household-based contact tracing model, we explore the probability that simulated epidemics that begin with a single infection go extinct over a 2-year period (assuming no additional importations), and the time that it takes them to do so. Because the level of physical distancing has a strong effect on the course of the epidemic, we show the relationship between epidemic end states and extinction times for 0- 90% reductions in global contacts with one starting infection and with 100 starting infections. If a branching process model of an epidemic experiences sufficient growth, it will never go extinct since it does not consider depletion of global susceptible contacts. As a result, simulated epidemics are stopped once they hit 5000 active infections, as the probability of extinction is effectively zero at this point and exponential growth is achieved. Some parameters are fixed as described in Table 1 and others were varied as described in Table 2. Additionally, we evaluate the extinction times for scenarios A and E of social distancing as described in Table 4 and 5.

The ‘non-complex’ NHS Test and Trace system uses individual-based tracing with tracing initiated on test results^56^. To assess the potential for improving the effectiveness of contact tracing using backwards tracing, we explore the relationship between the growth rate and the number of days prior to symptom onset over which contacts are traced using the individual-based tracing model described previously. This model includes household structure that impacts on infection dynamics, but only the contacts of individuals who test positive are traced.

We consider both individual level and household level contact tracing with and without a tracing app, and also consider a decay in contact recall as the number of days back to trace increases, a 10% daily decrease in probability of successfully recalling a contact. If an individual reports they suspect infection, then their household is quarantined. Testing is ordered for the individual, and anyone else in the household who has had symptom onset. Upon an individual receiving a positive test result, contact tracing is initiated for contacts that occurred up to X days prior to symptom onset, and up to 7 days post symptom onset, where X is the number of days prior to symptom onset over which contacts are traced and a parameter to be varied. We assumed an unvarying set of parameter values (as shown in Table 2) to explore the relationship between growth rates and the number of days back over which to trace.

### Concurrent Global Contact Reductions

While useful to demonstrate the effect of contact tracing in combination with physical distancing, uniform reductions in global social contacts is not inherently an interpretable scenario; the assumption that all households will perform physical distancing equally is expected to be violated in practice. As such, we consider assumed scenarios of different lockdown relaxations by conditioning the reduction in global contacts upon household size, relative to the mean number of global contacts by household size that was estimated in the POLYMOD survey^42^. The lockdown scenario was parameterised using CoMix data from the lockdown period from end March - end April 2020^23^.

The UK government plan to relax lockdown consisted of three stages^57^, starting with an increase in exercise allowed per day, followed by a phased return of children to schools and opening non-essential retail, followed by places of worship, leisure facilities and hospitality. We loosely based our assumed relaxation scenarios around this plan, though do not model household composition explicitly (Tables 3 and 4).

We ran 100 simulations for each relaxation scenario, varying all strategies and parameters across their priors. Adherence and uptake of quarantine was assumed to be perfect, and testing was not required for contact tracing to be initiated. The resulting distribution of the growth rates can be seen in Figure 6 and the end states of the simulations are described in Table 5.

## Results

### Effects of household structure and tracing on growth rates

In models with household structure (Figure 4B and 4C), the decline in growth rates associated with higher global contact reductions was less steep compared to epidemics with no household structure (Figure 4A). A reduction in global contacts is a smaller proportion of total contacts when household structure is modelled explicitly, because there are no local contacts when there is no household structure. To achieve a growth rate of zero, a higher percentage global contact reduction is required in models that include household structure (approximately 70%, no tracing) compared to those that do not (approximately 65%, no tracing).

**Figure 4:**
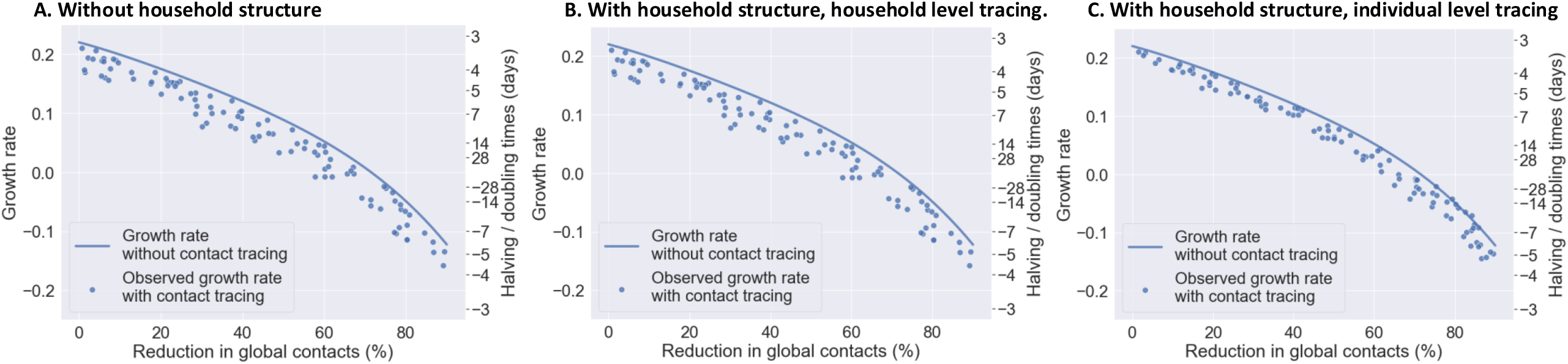
The effect on growth rates of contact tracing, quarantine and case isolation compared to case isolation only: the role of household structure and household-based contact tracing. Negative values on the doubling time axis imply it is a halving time and the epidemic is decreasing for these values. The growth rate without contact tracing and only case isolation (solid line) was derived by simulation of the branching process without contact tracing. Simulated epidemics with contact tracing are points and reflect both uncertainty in data-driven parameterisation and a range of plausible contact tracing parameter values (see Tables 1 and 2).

There was a greater variability in the outcomes of epidemics from models with household structure compared to the model with no household structure, but the potential gains in controlling the epidemics were greater when household-based tracing was used, for the same range of parameter values. The effects of tracing on growth rates was more sensitive to the case discovery probability in the model with household structure and tracing (0.0110 compared to 0.0078 for a 10% increase in the probability of infection discovery). However, when considering a model with household structure but individual-based tracing, the effects of contact tracing on the growth rate appeared lower (an approximate 0.02 reduction in the growth rate day^-1^ compared to epidemics without tracing).

### Growth rates for a range of household-based contact tracing strategies

In a baseline case where individuals with symptoms and their households self-isolate and their household members quarantine as per UK policy, but there is no contact tracing, the unconstrained growth rate drops below 0 when there is approximately at least a 70% reduction in global contact rates (Figure 5). Household-based contact tracing initiated on symptoms report, varying parameter values across those given in Table 2 (including two-step tracing, unlike in the household comparison simulations above), reduced the growth rate of simulated epidemics by approximately 0.05 day^-1^ compared to baseline across the range of global contact reductions. This controlled the epidemic at global contact reductions of less than 50% for some simulated epidemics but not all.

**Figure 5:**
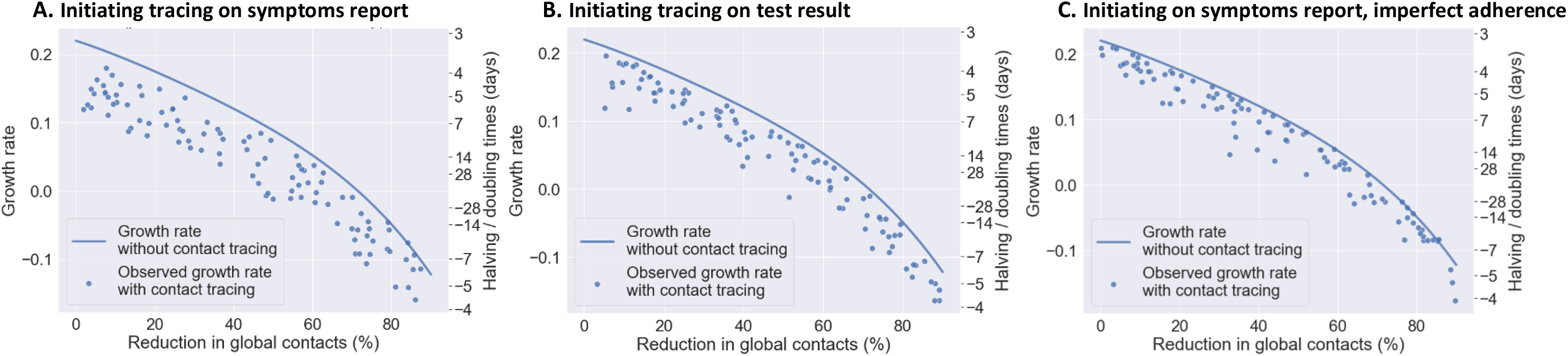
The effect on growth rates of contact tracing, quarantine and case isolation compared to case isolation only when contact tracing is initiated upon A. case symptoms; B. case positive test result; and C. case symptoms, with imperfect adherence. Negative values on the doubling time axis imply it is a halving time and the epidemic is decreasing for these values. The growth rate without contact tracing and only case isolation (solid line) was derived by simulation of the branching process without contact tracing. Simulated epidemics with contact tracing are points and reflect both uncertainty in data-driven parameterisation and a range of plausible contact tracing parameter values (see Tables 1 and 2).

For each one day increase in mean testing delay, the growth rate was associated with an increase of 0.0138 (95% CI 0.009-0.018, Table 7) when the mean testing delay was varied across the range of 1.5 to 2.5. This represents a substantial decrease in contact tracing efficacy as testing delays increase.

**Table 6:** Effects of contact reductions and contact tracing parameters on growth rates across models with and without household structure and household-based contact tracing.

| Parameter | Without household structure | With household structure, household level tracing | With household structure, individual level tracing |
| --- | --- | --- | --- |
| Intercept<br>(instantaneous growth rate) | 0.2400<br>(0.220, 0.260) | 0.2402<br>(0.224, 0.256) | 0.2353<br>(0.220, 0.251) |
| Reduction in global contacts (per 10% reduction in global contacts) | -0.0212<br>(-0.031, -0.011) | -0.0132<br>(-0.021, -0.005) | -0.0151<br>(-0.024, -0.006) |
| (Reduction in global contacts)<sup>2</sup> (per 10% reduction in global contacts) | -0.0040<br>(-0.008, 0.000) | -0.0054<br>(-0.009, -0.002) | -0.0036<br>(-0.007, 0.000) |
| (Reduction in global contacts)<sup>3</sup> (per 10% reduction in global contacts) | 0.0008<br>(1.15e-5, 0.002) | 0.0008<br>(0.000, 0.001) | 0.0005<br>(-0.000, 0.001) |
| (Reduction in global contacts)<sup>4</sup> (per 10% reduction in global contacts) | 7.42e-5<br>(-0.000, -3.32e-5) | 5.78e-5<br>(9.5e-5, 2.45e-5) | -3.865e-5<br>(-7.15e-5, 5.78e-6) |
| (Probability of having the tracing app)<sup>2</sup> (per 0.1 increase in probability) | -0.0004<br>(-0.001, -0.000) | -0.0001<br>(-0.000, 2.36e-5) | 1.716e-5<br>(-0.000, 0.000) |
| Probability that a contact made is successfully traced (per 0.1 increase in probability) | -0.0054<br>(-0.007, -0.004) | -0.0053<br>(-0.007, -0.004) | -0.0024<br>(-0.004, 0.001) |
| Mean contact tracing delay (per one day increase) | 0.0101<br>(0.006, 0.015) | 0.0055<br>(0.001, 0.010) | 0.0014<br>(-0.002, 0.005) |
| Mean testing delay (per one day increase) | 0.0094<br>(0.005, 0.014) | 0.0097<br>(0.006, 0.013) | 0.0060<br>(0.002, 0.010) |
| Infection discovery probability (per 0.1 increase in probability) | -0.0078<br>(-0.009, -0.007) | -0.0111<br>(-0.012, -0.010) | -0.0091<br>(-0.010, -0.008) |

*Results of log-linear regression. We performed 100 simulations for each model, with 5000 starting infections and estimated the growth rates using days 15 - 25 of the simulation and the process had become well mixed. All simulations included testing cases before tracing their contacts. There is no interpretation of the intercept because we do not simulate scenarios where there is no contact tracing.*

**Table 7:** Effects of contact reductions and contact tracing parameters on growth rates when contact tracing is initiated upon A. case symptoms; B. case positive test result; and C. case symptoms, with imperfect adherence.

|  | A. Initiating tracing on symptoms report | B. Initiating tracing on test result | C. Initiating on symptoms report, imperfect adherence |
| --- | --- | --- | --- |
| Intercept<br>(Instantaneous growth rate) | 0.2891<br>(0.270, 0.308) | 0.2203<br>(0.193, 0.247) | 0.3311<br>(0.310, 0.352) |
| Two step tracing at the household level (True) | -0.0067<br>(-0.009, -0.004) | -0.0106<br>(-0.013, -0.008) | -0.0007<br>(-0.004, 0.002) |
| Reduction in global contacts (per 10% reduction in contacts) | -0.0214<br>(-0.032, -0.010) | 0.0020<br>(-0.015, 0.019) | -0.0227<br>(-0.034, -0.011) |
| (Reduction in global contacts)^2 (per 10% reduction in contacts) | -0.0023<br>(-0.007, 0.003) | -0.0103<br>(-0.017, -0.004) | -0.0023<br>(-0.008, 0.003) |
| (Reduction in global contacts)^3 (per 10% reduction in contacts) | 0.0004<br>(>-0.001, 0.001) | 0.0015<br>(4.73e-4, 0.003) | 0.0005<br>(-3.46e-4, 0.001) |
| (Reduction in global contacts)^4 (per 10% reduction in contacts) | -0.408e-5<br>(-8.72e-5, 5.63e-6) | -9.36e-5<br>(-1.48e-4, -3.91e-5) | -5.11e-5<br>(-9.92e-5, -2.91e-6) |
| (Probability of having the tracing app)^2 (per 0.1 increase in probability) | -0.0004<br>(-0.001, -1.96e-4) | -0.0003<br>(-4.70e-4, -0.009) | -0.0001<br>(-3.55e-4, 1.02e-4) |
| Probability that a contact made is successfully traced (per 0.1 increase in probability) | -0.0106<br>(-0.012, -0.009) | -0.0050<br>(-0.070, -0.030) | -0.0059<br>(-0.008, -0.004) |
| Mean contact tracing delay (per one day increase) | 0.0152<br>(0.011, 0.019) | 0.0104<br>(0.005, 0.015) | 0.0067<br>(0.002, 0.012) |
| Infection discovery probability<br>(per 0.1 increase in probability) | -0.0227<br>(-0.028, -0.018) | -0.0205<br>(-0.026, -0.016) | -0.0073<br>(-0.013, -0.002) |
| (Infection discovery probability)<sup>2</sup><br>(per 0.1 increase in probability) | 0.0009<br>(1.63e-4, 0.002) | 0.0013<br>(4.81e-4, 0.002) | 0.0001<br>(-0.001, 0.001) |
| Mean testing delay<br>(per one day increase) | N/A | 0.0138<br>(0.009, 0.018) | N/A |
| Probability a household will take up isolation (per 0.1 increase in probability) | N/A | N/A | -0.0098<br>(-0.0011, -0.009) |

*Results from log-linear regression. Parameters were fixed or varied as described in tables 1 and 2. We performed 100 simulations for each model, with 5000 starting infections and estimated the growth rates using days 15 - 25 of the simulation and the process had become well mixed. Note that the intercept has no interpretation because we do not simulate scenarios with no contact tracing here.*

**Table 8:** End states of simulated epidemics with 1 starting infection across the assumed scenarios of physical distancing relaxation.

| Scenario | % Epidemics that did not reproduce | % Epidemics go extinct within 730 days | % Epidemics that hit 5000 active infections | % Epidemics that hit 2 years without going extinct |
| --- | --- | --- | --- | --- |
| A | 45.8 | 54.2 | 0 | 0 |
| B | 39.8 | 60.2 | 0 | 0 |
| C | 35.3 | 64.0 | 0.3 | 0.4 |
| D | 30.4 | 61.6 | 7.8 | 0.2 |
| E | 20.6 | 36.1 | 43.3 | 0 |

*Lockdown relaxation scenarios described in Tables 4 and 5, with scenario A representing a small increase in school and workplace contacts, and scenario E representing a larger increase in work contacts, resumption of school contact and resumption of most leisure contacts*

Following global contact reductions, the parameter with the greatest effect on epidemic growth across scenarios was the probability that infections would be discovered (Table 7). This is perhaps unsurprising since all other tracing parameters are dependent upon this.

Two step tracing had a greater effect when tracing was initiated on a positive test result rather than on symptoms (Table 7). Fundamentally two step tracing is a strategy that improves the speed of the contact tracing process by tracing contacts of contacts of an infected case, as opposed to waiting until a first generation contact develops symptoms and tests positive. As such, the improvement from two step tracing is greater when it is used to offset the slowdown caused by testing every individual before propagating contact tracing. We assume that it does not affect the probability that a contact is successfully traced, though if a model of recall is considered there can be interactions with the probability a contact is recalled, since first generation contacts would be asked to recall their contacts earlier. An effect of two-step tracing was not observed in the model including non-uptake and waning adherence to isolation and quarantine: its slight gains appear to have been eroded by the non-adherence effects. Non-uptake of isolation or quarantine was associated with higher growth rates, though a waning effect on adherence to isolation or quarantining (ie, leaving early) was not.

The tracing delay was less important when considering tracing on test receipt rather than on symptoms and when considering non-uptake and non-adherence to quarantine. The tracing delay occurs relatively late in the transmission process, after the time until case discovery (given that not all are detected) and after waiting for testing, so the relative gains or losses that can be seen at this stage in the process are fewer.

### Backwards Contact Tracing and Recall

We consider the role that implementing ‘backwards tracing’ might play in improving the effectiveness of England’s Test and Trace policy, by varying the number of days prior to symptom onset over which to trace contacts.

We find that increasing the number of days prior to symptom onset over which contact tracing is performed improves the efficacy in reducing epidemic growth rate (Figure 6A), especially when app uptake is high (50%, Figure 6C) resulting in more digital contact tracing which has no contact tracing delay and more contacts successfully traced. There is a linear decrease in the growth rate up until around 8 to 10 days, where there are no more gains to be made.

**Figure 6:**
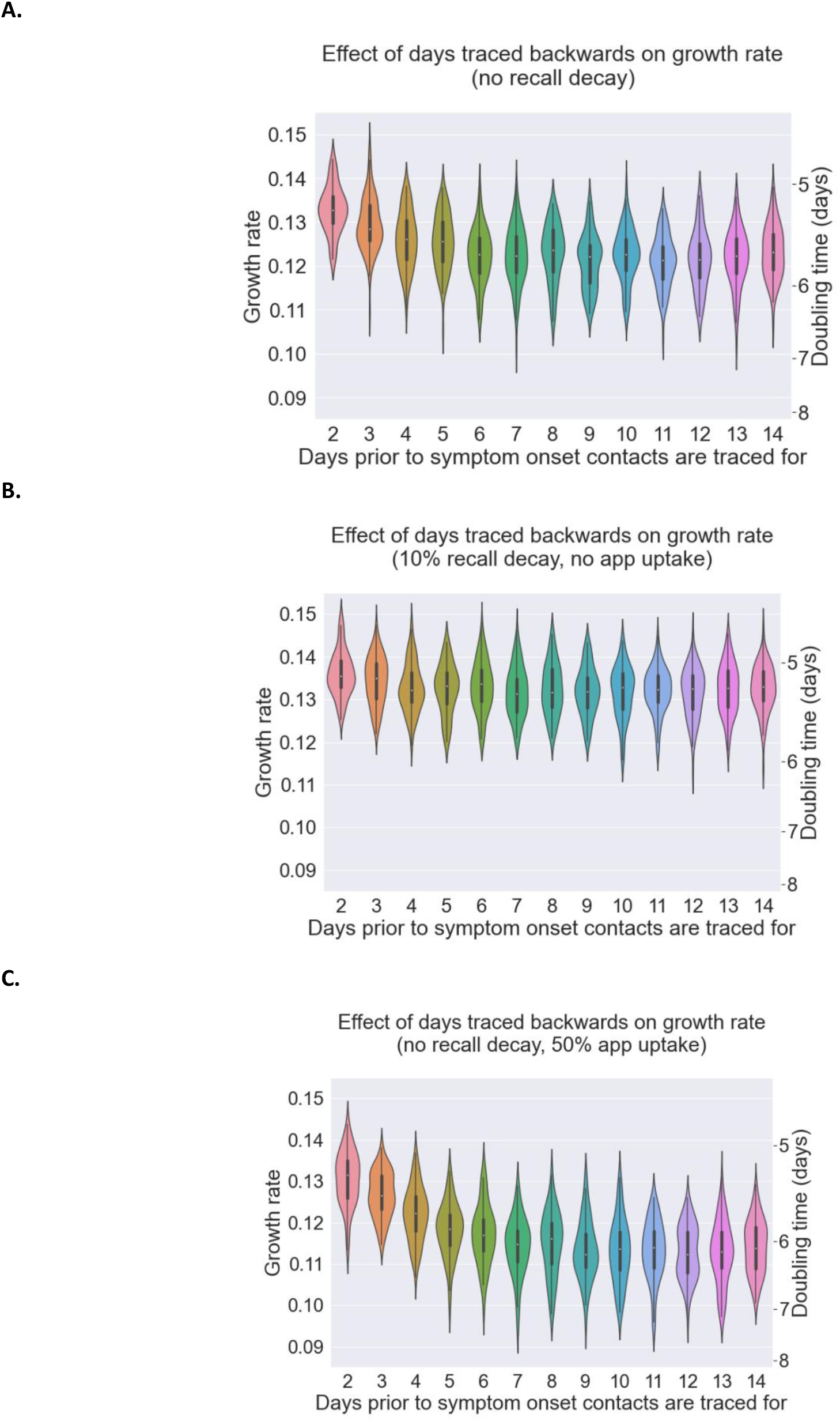
The effect of days traced back from case symptom onset on growth rate considering A. no contact recall decay, B. daily 10% contact recall decay and C. no recall delay and 50% tracing app uptake.

However, identified cases might struggle to remember contacts that they have had further back in the past. We find that including a daily 10% reduction in the probability of recalling a contact in the model erodes the gains of backwards contact tracing, almost completely, such that there is little difference in the growth rate of the epidemic according to the number of days pre-symptom onset a case’s contacts are traced, assuming that there is a 10% daily drop in the probability that a contact is recalled by a case since the day of last contact, Figures 6B. The overall reduction on the growth rate of the epidemic is limited. For the index case in a contact tracing chain to have occurred, the index case has had symptom onset, a symptom reporting delay and a possible testing delay before the backwards tracing attempt to the infector is initiated, this already would lead to significant decay in the ability to recall. The infector will be reached after a further contact tracing delay and will then have to recall their contacts that occurred at around the time they contacted the index node. As a result, the recall decay significantly impacts the probability that backwards then forwards tracing can occur.

### Lockdown exit scenarios

We plot the posterior distribution of the growth rates under the assumed scenarios (described in Tables 4 and 5) in Figure 7. With household-based contact tracing and global contact patterns analogous to scenarios A and B, (small increases in school and workplace contacts for A, an additional 10% increase in leisure contacts for B), the growth rate for all simulations remained below 0. However, for Scenarios C and D, the results for positive or negative growth were mixed, with nearly all simulated epidemics with contact levels analogous to Scenario E finding positive growth of the epidemic.

**Figure 7:**
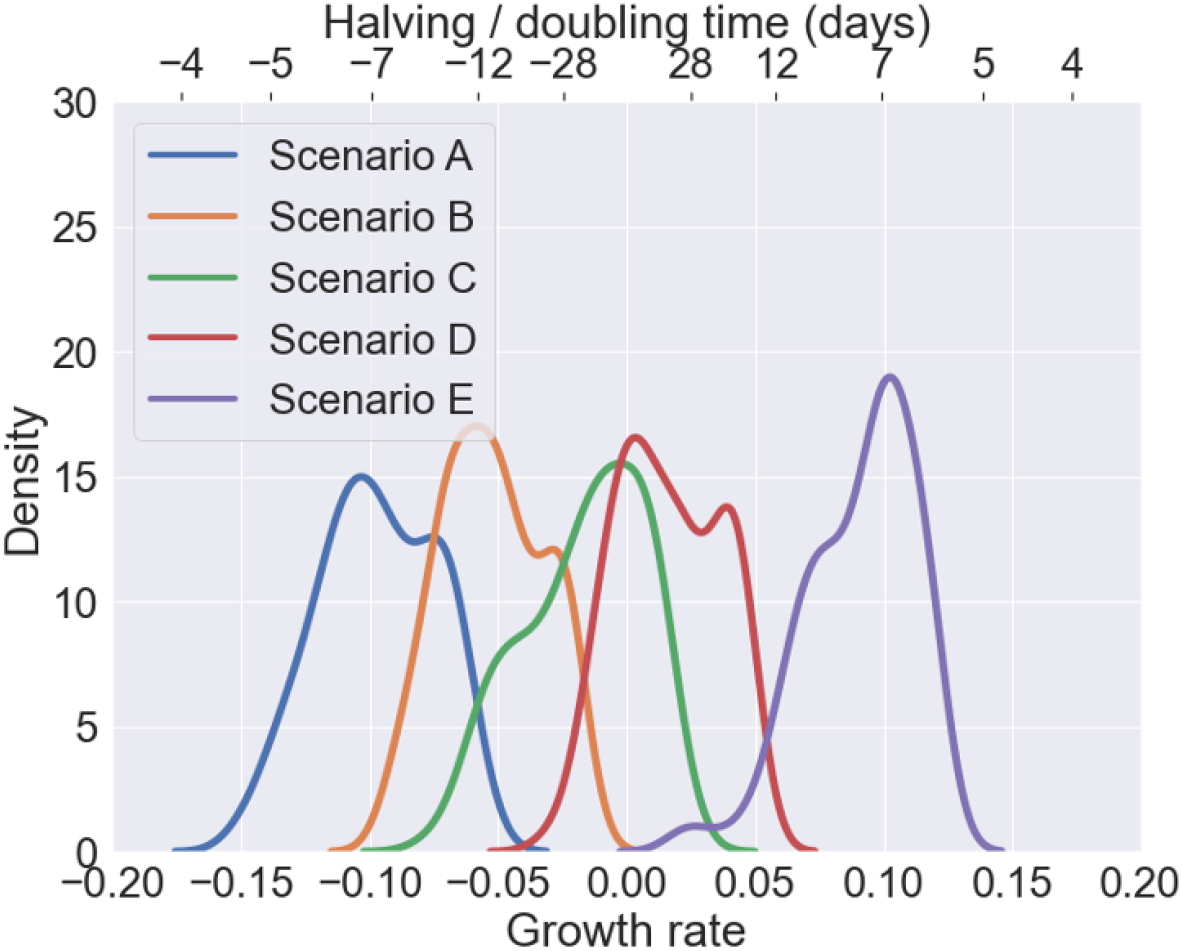
Observed growth rate distributions of the epidemic growth rate under the different lockdown relaxation scenarios described in Tables 4 and 5, with scenario A representing a small increase in school and workplace contacts, and scenario E representing a larger increase in work contacts, resumption of school contact and resumption of most leisure contact. Negative values on the doubling time axis imply it is a halving time.

### Extinction Times

We simulated epidemics of a timeframe of 730 days to estimate the proportion of epidemics that go extinct and the time taken to do so, varying contact tracing parameters as in Table 2.

We define several possible end states for an epidemic; ‘No reproductions’, where the first generation produces no offspring; ‘Hit 5000 active infections’, at which point we assume the probability of extinction is 0 due to the size of the epidemic, and the simulation is stopped; ‘Hit 2 year time limit’, where we stop simulating epidemics that were not extinct after 2 years nor hit 5000 active infections; and finally ‘Extinct’ where the first generation produces some offspring, but the number of active infections then drops to 0 and no more infection events occur. These ‘end states’ depend strongly on the global contact reductions (Figure 8). For a single infection, approximately half of epidemics go extinct if the global contact reduction is at least 50%. For 100 starting infections approximately half of simulated epidemics go extinct at a contact reduction of approximately 65%, with variation in the end states occurring between 50 and 70%. That is, below a 50% global contact reduction, contact tracing and quarantine does not bring the epidemic under control (which can also be seen on the growth rate plots, Figure 5A). In the 50-70% range, the mean and variance in extinction times reduces with greater contact reductions.

**Figure 8:**
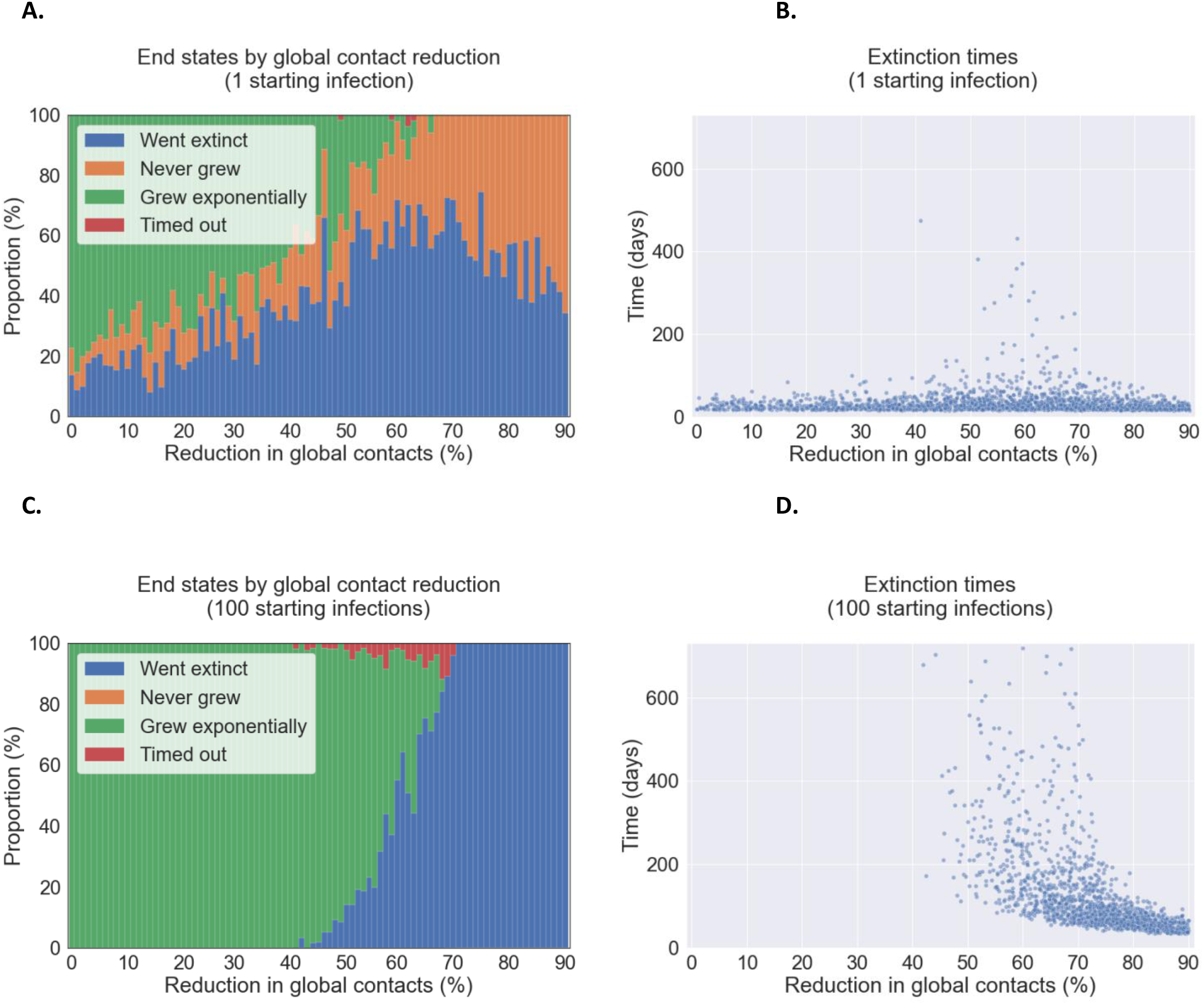
End states and extinction times for simulated epidemics.

**Figure 9:**
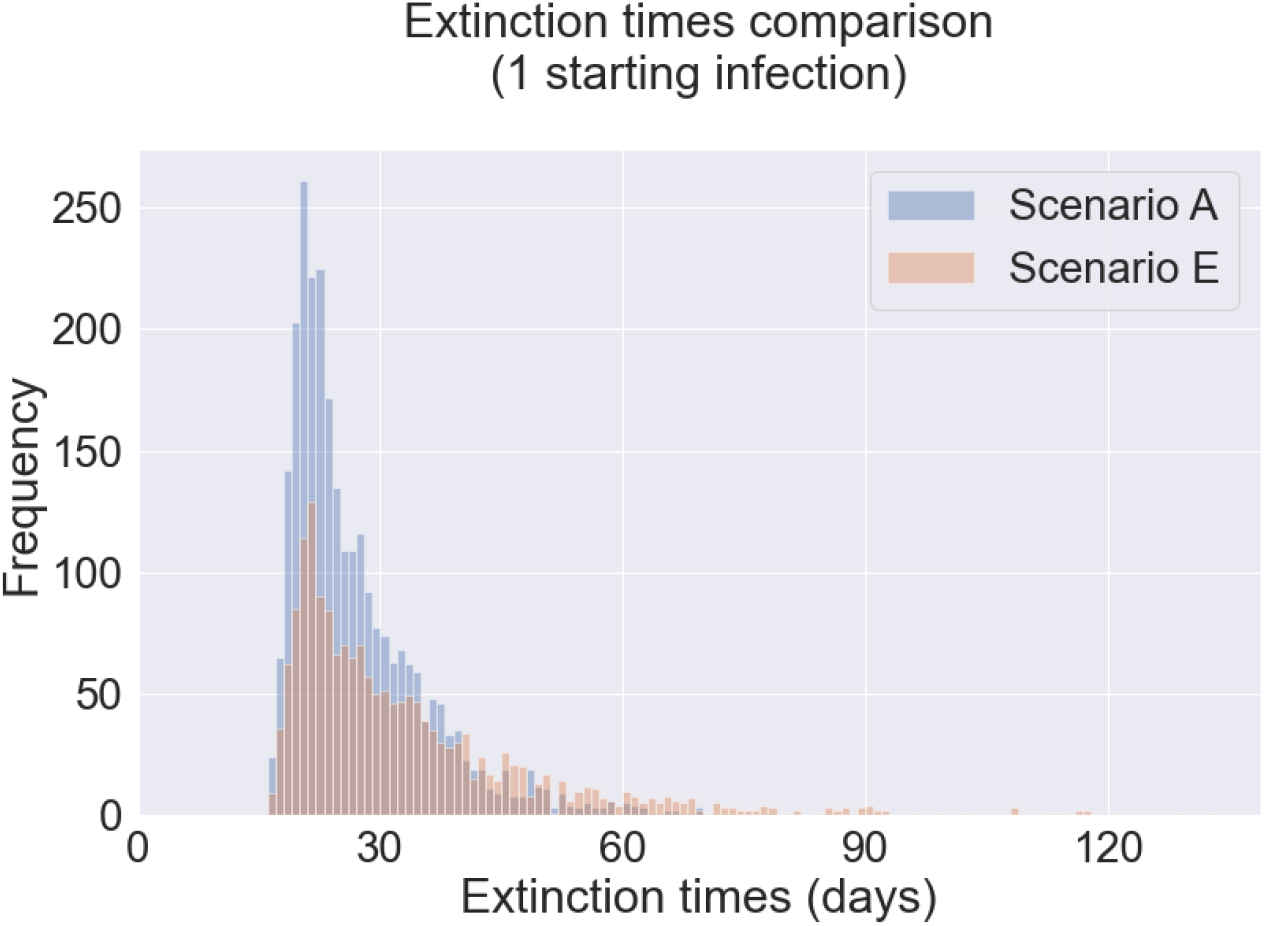
Extinction times of epidemics started with a single infection that reproduced, for lockdown relaxation scenarios A and E. Lockdown relaxation scenarios are described in Table 5, with scenario A representing a small increase in school and workplace contacts, and scenario E representing a larger increase in work contacts, resumption of school contact and resumption of most leisure contact. All other parameters varied as described in Table 2.

Considering lockdown exit scenarios linked to increases in work, school and leisure contacts, from Scenario A to E, we found that the likelihood of an initial infection causing secondary infections increased (Table 6), and that once this occurred, there were more epidemics that could not be controlled. Among epidemics that were controlled, there were more simulated epidemics which took longer to go extinct in Scenario E compared to A (Figure 7). This highlights the long time frame over which interventions must be kept in place.

When we considered epidemics with no physical distancing, the probability of epidemic extinction arising from a single initial infected individual was much lower, even given contact tracing with 100% adherence to quarantine, (not shown here).

## Discussion

We find that implementing a household contact tracing and isolation policy could contribute to controlling the SARS-CoV-2 epidemic if lockdown levels of physical distancing are partially relaxed, but not if they are relaxed completely. In our household structured branching process model, none of our strategies to improve contact tracing would prevent the re-emergence of epidemic growth once global contact reductions relative to pre-pandemic times are reduced by less than approximately 40%. We found that contact tracing and isolation could contribute to reducing epidemic growth for contact reductions above this level but that the probability that cases are discovered is key. We have explored strategies that could be used to make contact tracing more effective. Notably, household-based tracing was more effective in reducing the growth rate of epidemics than individual-based tracing, though this and other strategies need to be considered in terms of the increased number of individuals who would need to be traced and go into isolation. The extinction times of some simulated epidemics could be potentially very long, which underlines the need to account for sustainability of interventions over time. Consideration should be given to our findings that effectiveness gains could be eroded both by implementation challenges, such as contact recall difficulties in the case of backwards tracing, and by non-uptake and waning adherence to isolation and quarantine.

Countries that have suppressed the number of SARS-CoV-2 cases have done so with a combination of policies, including contact tracing and varied levels of suggested and enforced physical distancing policies. Our findings that contact tracing and isolation are unlikely to be effective on their own in suppressing the SARS-CoV-2 epidemic are consistent with those of other models^22,58,59^. The contribution of contact tracing and isolation in our model was most significant when global contact reductions were 40-70% of their usual levels, under which growth was not suppressed and over which very few epidemics took off (contingent on the parameters of transmission that we chose). Further evidence on the transmissibility amongst different groups, types of contacts and in different settings will allow a more nuanced interpretation of which physical distancing policies are most important to retain.

Following the global contact rate, the probability that a case is identified was the next most important model parameter to the effectiveness of contact tracing, isolation and quarantine. In our model, this parameter reflects a combination of biological factors (e.g. proportion of asymptomatic and subclinical infections), testing policies (who is tested and when in their infection), and test characteristics (sensitivity and specificity). Estimating the proportion of cases who are discovered overall is challenging; studies in other European countries indicate that the overall proportion of cases who are discovered is low (20% in Spain, 10% in France post-lockdown^60,61^). Our findings suggest that even with a contact tracing, isolation and quarantine system in place, additional case discovery methods would be beneficial, for instance frequent screening of high-risk groups. This is analogous to the approaches taken for the control of sexually transmitted infections, which utilises both screening of high-transmission risk groups and tracing (partner notification), but requires an understanding of the links between ‘high’ and ‘low’ risk individuals and how their definition changes over the course of the epidemic^62^.

Once the number of global contacts and the proportion of cases discovered are in a range in which contact tracing and isolation could hypothetically control the epidemic, the speed of the process relative to the speed of transmission - fast in the case of SARS-CoV-2 - is important. Strategies to either minimise or eliminate delays, including symptom-initiated tracing rather than waiting for test results, and smartphone app tracing were more effective in suppressing epidemic growth compared to manual tracing (when uptake of the hypothetical app was high, as others have found^63^). Other strategies to ‘get ahead’ of transmission, including earlier isolation of contacts via two-step contact tracing and isolation or concurrently isolating a contact’s whole household, also reduced growth rates but less significantly so. Backwards tracing improved effectiveness up to tracing 8-10 days prior to symptom onset assuming no change in the probability of successfully tracing contacts, after which presumably the time delay incurred before tracing ‘forwards’ again on previously missed transmission branches becomes too great to ‘catch-up’.

However, for each of the strategies that could theoretically improve the effectiveness of contact tracing, there are implementation challenges that could erode their effectiveness. When considering a possible decrease in successfully tracing contacts back in time to enable backwards tracing, we found that a plausible ‘worst case’ assumption in declining recall of contacts could almost cancel out any gains made. Many outbreaks occur in settings with specific environmental conditions, so implementing backwards tracing by asking individuals where they have been, instead of who they met, could help facilitate the discovery of clusters of infections.

If uptake of isolation policies is less than 100% and if adherence to isolation wanes over the period of isolation, during which time household transmission could continue, we found that effectiveness in suppressing epidemic growth degraded. Monitoring of adherence and policies to enable and support people to take up and adhere to testing, tracing and isolation are crucial. In our analysis, the time it would theoretically take to eliminate SARS-CoV-2, could be in the order of months or years given slight relaxations of lockdown. These long theoretical extinction time estimates assume no further importation of cases into the population, which is unrealistic in practice, suggesting that control policies would be required over a long timeframe.

### Strengths and Limitations

Our model’s household structure enables more realistic transmission dynamics and explicit modelling of household quarantines and isolation and the strategic options these create. Interactions between physical distancing and household structure are important to consider - a 90% reduction in global contacts does not necessarily correspond to a 90% reduction in transmission, as the within-household epidemics will continue to spread. When there are high levels of global contact reductions, individuals are spending more time within the household which may result in increased levels of local transmission which we have not modelled here. We found that models that do not include households risk under-estimating the potential impact of a household-based tracing strategy or overestimating the effectiveness of contact tracing when an individual-based tracing strategy is used in a real population with households. However, we do not include other important clustering in our model that would connect households, such as workplaces or schools, which are important to consider as children return to school and more workers are encouraged to stop working from home.

Our model does not account for global susceptible depletion. In practice there will be many individuals who repeat the same global contacts each day, such as office workers who should experience susceptible depletion in the number of colleagues they can infect^64^. Further, we expect individuals in the same household to share some social contacts, which also results in increased global susceptible depletion, and fewer opportunities to infect globally. As the epidemic continues, modelling a level of immunity will be required. In the UK, ONS has estimated seroprevalence from antibody testing on blood donors at approximately 6% in England (as of August 23, 2020), with regional variations^18^. It is also not yet known how durable the immune response to a repeat exposure will be, and how this might vary by severity of the initial infection and other factors. Levels of immunity will continue to be heterogenous by region and sociodemographic factors, which needs to be accounted for. Contact network effects would further come into play, as elements of the population who make a large number of contacts on a regular basis will have an increased likelihood of immunity. Our model is currently limited to households, and other settings such as schools and workplaces must be considered. We have not explicitly modelled ‘support bubbles’, whereby households of single individuals have been allowed to function as one larger household under physical distancing restrictions^46–48^. We do not model individual-level heterogeneity, such as age, specific vulnerabilities to severe infection or characteristics associated with increased exposure and transmission, and the relationship to household structure. Network effects are not included in these models, which could mean we are potentially underestimating the effectiveness of contact tracing, which preferentially removes individuals with many contacts (those with ‘high-degree’)^10^, though this might be subject to other epidemic characteristics and contact tracing performance^36,53,65^. Our framework will allow for the addition of other settings, attributes and network structures going forward.

Relatedly, we do not explicitly model the ‘costs’ to different contact tracing and isolation strategies and choices, including the number of people required to isolate per identified case. This will vary according to contact patterns in the population at a given point in time, and the impact on individuals and society for instance via isolation of key workers, will also vary. The risk of app tracing to identifying a large number of non-epidemiologically relevant contacts is another potential problem to consider. Previous studies of quarantine orders vary widely in their findings as to adherence and patterns over time^25^. We also do not model indirect benefits to reducing transmission that contact tracing could bring, such as improved surveillance and understanding as to transmission patterns that could enable better timed and targeted interventions.

We conclude that while there are strategies to improve the effectiveness of contact tracing, isolation and quarantine on epidemic growth, contact tracing and isolation as we have modelled it will not be effective in suppressing epidemic growth on its own, given current understanding about transmissibility of the virus without continued reductions in social contacts. To be effective, contact tracing and isolation will need to be performed with minimal delays and a high degree of accuracy. Further, it will be important to consider support to uptake and adherence to policies, and to better understand potential trade-offs between strategies that reduce growth but which might have a negative effect upon adherence, such as smartphone app based tracing. Thought needs to be paid to practical implementations in order to gain the theoretical effect. It is likely that as more about transmission settings and dynamics are understood, testing, contact tracing and isolation can continue to be refined and will remain an important part of a targeted approach to the control of SARS-CoV-2.

## Data Availability

This study did not collect any data. All code used is given at the repository URL below.

https://github.com/martyn1fyles/HouseholdContactTracing

## Acknowledgements

EF is funded by the MRC (grant MR/S020462/1). MF is supported by The Alan Turing Institute under the EPSRC grant EP/N510129/1. LP, and CO are funded by the Wellcome Trust and the Royal Society (grant 202562/Z/16/Z). TH is supported by the Royal Society (Grant Number INF/R2/180067) and Alan Turing Institute for Data Science and Artificial Intelligence. IH is supported by the National Institute for Health Research Health Protection Research Unit (NIHR HPRU) in Emergency Preparedness and Response and the National Institute for Health Research Policy Research Programme in Operational Research (OPERA) and Alan Turing Institute for Data Science and Artificial Intelligence. TW is supported by grants from: the Wellcome Trust, UK (209075/Z/17/Z); the Medical Research Council, Department for International Development, and Wellcome Trust (Joint Global Health Trials, MR/V004832/1), the Academy of Medical Sciences, UK; and the Swedish Health Research Council, Sweden.

