## Supplementary material for "Using a household structured branching process to analyse contact tracing in the SARS-CoV-2 pandemic"

#### 1 Introduction

In this document we discuss the underlying mathematical framework, model assumptions, distributions, processes and analysis of our model. In particular, we use a network framework in which we think of the individuals who have ever been infected by time $t$ as forming a set of *nodes* (or *vertices*), $\mathcal{V}_t$. If there has been a transmission event from the individual $v \in \mathcal{V}_t$ to the individual $v' \in \mathcal{V}_t$ by time $t$ then $(v, v') \in \mathcal{E}_t \subset \mathcal{V}_t \times \mathcal{V}_t$ and we call $\mathcal{E}_t$ the set of *edges*. In this way, the epidemic can be represented by a time-indexed network $\mathcal{G}_t = (\mathcal{V}_t, \mathcal{E}_t)$.

#### 2 Branching Processes

Branching processes are a well established tool for studying epidemics, and offer a mathematical framework for understanding the initial exponential growth, stochastic variability around the expected behaviour, extinction times and extinction probabilities. Further, it is possible to model a contact tracing process as a type of *super-infection* along the branching process as an approximation of contact tracing interventions [4]. We model the SARS-CoV-2 epidemic using a branching process with two levels – individuals and households – for several reasons. Firstly, within-household transmission is important for directly transmitted infectious diseases such as influenza [3] and SARS-CoV-2 [8], and by incorporating a model of household structure we develop a better approximation of the epidemic. Secondly, when contact tracing discovers an infection in a household, this should lead to all household members being rapidly informed of their exposure from their housemate and going into quarantine. The ability of a contact tracing process to reduce the spread of the virus can therefore be improved if the household structure is leveraged. Finally, there are interesting effects that occur at the household level that cannot be be ignored, such as individuals in a larger household typically making more contacts pre-lockdown, or smaller households having less ability to socially distance, for example in a larger household one person can do the shopping for several people.

Pellis et al. [11] used a multitype branching process to define and calculate an epidemic threshold value $R^*$ for household models. We form the basis of our model using their multitype branching process which approximates an epidemic in a population segmented in households of varying sizes with an assumption of homogenous mixing between the households. Briefly, in a multitype branching process each node is one of a finite number of types $d$. Let $M = [m_{ij}]_{i,j \in 1 \ldots d}$ be the mean offspring matrix where $m_{ij}$ is the expected number of type-$j$ children of a type-$i$ node; then $R^*$ is defined to be the largest eigenvalue of $M$. The type of a node is defined by its household size and which generation of the local epidemic it belongs to. A node's offspring must either be a generation 0 infection in a new household, or in the following generation of the within-household epidemic. A simplifying assumption is made when deriving the household branching process where the virus is always introduced into a fully susceptible household, as such this represents a worst case scenario for the epidemic - introduction of the virus into a household where there is some immunity would lead to fewer secondary infections when compared to introduction of the virus into a fully susceptible household.

### 3 Contact Model

We must assume an underlying mechanism and model for the spread of the infection. Given that we aim to analyse contact tracing in combination with social distancing measures, it makes sense to use social contact distributions to form the basis of the model. Each day a node makes a random number of contacts *locally* (within household contacts) and *globally* (outside household contacts) which can provide opportunities for the virus to spread. If the node making a contact is infected, then a the contact carries a probability of spreading the infection, conditional upon the node's infectious age (the amount of time that has passed since they were infected). Later on, we model social distancing measures by reducing the number of contacts that occur.

The POLYMOD survey [9] collected data on the number of social contacts made by individuals in a day, and we use this to provide parameters for the underlying contact distribution for a pre-lockdown time period. POLYMOD performed social contact surveys in several European countries, and we make use of the findings for the United Kingdom. Nodes are considered to be identical in nature apart from the size of the household they belong to, there are no subclasses of nodes such as children or adults. Let $C_h$ be the number of contacts made by an individual in a household of size $h$, we assume that $C_h \sim \text{OverdispersedNegBin}(\mu_h, \phi)$. The probability density function of the overdispersed negative binomial is given by

$$\mathbb{P}(C_h = c) = \frac{\Gamma(c + 1/\phi)}{\Gamma(c+1)\Gamma(1/\phi)} \left(\frac{1}{1+\phi\mu_h}\right)^{1/\phi} \left(\frac{\phi\mu_h}{1+\phi\mu_h}\right)^{c}. \tag{1}$$

In this parameterisation, $\mathbb{E}(C_h) = \mu_h$ and $\text{Var}(C_h) = \mu_h(1 + \phi\mu_h)$; we recover a Poisson distribution in the limit $\phi \to 0$ and larger values of $\phi$ correspond to greater overdispersion in the number of contacts made in a given day, which leads to overdispersion in the distribution of the network generated by the infection process. We estimate the following mean values of contacts made by a node, conditional upon their household size.

| **Parameter** | **Value** |
|---|---|
| $\mu_1$ | 7.2 |
| $\mu_2$ | 10.1 |
| $\mu_3$ | 11.4 |
| $\mu_4$ | 12.8 |
| $\mu_5$ | 14.5 |
| $\mu_6$ | 15.8 |
| $\phi$ | 0.32 |

Table 1: Contact Distribution Parameters

#### 3.1 Size Biased Household Distribution

When a global contact spread the infection outside of the household we have to draw the household size of the newly infected node. Let $\mathbb{P}(H = h)$ be the probability that a household selected uniformly at random from the population of households has size $h$, for $h \in 1, \ldots, h_{\max}$. If we begin by assuming that all nodes make the same number of global contacts independent of household size, then if we choose a node uniformly at random we draw a household size from the size biased distribution of households:

$$\pi(h) = \frac{h\mathbb{P}(H = h)}{\sum_h^{h_{\max}} h\mathbb{P}(H = h)}$$

This is to reflect the ability of each member of a household to import the virus into the household, and therefore larger households are more likely to import the infection than their smaller counterparts.

In our model the assumption that all nodes make the same number of contacts does not hold true, since nodes in larger households tend to make more contacts on average and as a result have a higher probability of being infected. As such when a node is infected we must draw $h$ from a size and mean contacts biased distribution (2), where $\mu_h^{global}$ is the mean number of global contacts that a member of a household of size $h$ makes.

$$\pi^*(h) = \frac{h\mu_h^{\text{global}}\mathbb{P}(H = h)}{\sum_{j=1}^{h_{\max}} j\mu_j^{\text{global}}\mathbb{P}(H = j)} \tag{2}$$

Suppose that a node in a household of size $h$ makes $c$ contacts in total. We need to divide these contacts into global and local contacts, $c = c_{local} + c_{global}$ subject to $c_{local} \leq h - 1$. Following the techniques of [13], we can estimate the probability of contacting a member of a household from Polymod. If a contact occurred in setting "at home", with frequency "daily" and the age of the contact matches the age of a household member, then we consider the contact to be a local contact. When we estimate local contact probabilities we assume homogeneous mixing within the household. We assume that there is a fixed probability of contacting a given household member each day, leading to a binomial distributed number of local contacts. Children are deliberately oversampled in Polymod, as they typically play an important role in epidemics, as such our estimates will be slightly biased.

| **Household Size** | $\mathbb{P}_h$**(pairwise local contact)** | **Sample Size** |
|---|---|---|
| 2 | 0.826 | 219 |
| 3 | 0.795 | 217 |
| 4 | 0.803 | 289 |
| 5 | 0.787 | 127 |
| 6 | 0.819 | 32 |

Table 2: Household member contact probabilities

Each day we draw $c \sim C_h$ and $c_{\text{local}} \sim \text{Bin}(h - 1, \mathbb{P}_h(\text{local contact}))$. We require that $c \geq c_{\text{local}}$, if an illegal combination occurs we redraw from both distributions until a legal combination occurs.

#### 3.2 Social distancing

Interventions to stop epidemics do not occur in isolation, but in combination with one another. Social distancing and lockdown measures have been employed across the world to slow the spread of SARS-CoV-2. It is highly unlikely that contact tracing alone will control the epidemic, instead we are interested in finding out what combination of intervention will suppress the epidemic. A key question is learning how much relaxations of lockdown is permitted by widescale contact tracing.

We begin by discussing a *uniform* social distancing measure, where the same reduction in global contacts is applied to all members of the population. This approximation allows for an easily interpretable results for different levels of social distancing. In practice we expect the level of social distancing performed by an individual to be partly conditioned upon their household size. We therefore consider a second model of social distancing, *household level* social distancing where the amount of social distancing is conditioned upon the size of an individuals household. While more realistic, household conditional social distancing is most interpretable when an underlying scenario is assumed.

Uniform social distancing is modelled by a Bernoulli thinning of a nodes attempted global contacts with parameter $\rho$. As such, if a node intended to make $c_{\text{global}} \sim C_{\text{global}}^{(h)}$ contacts that day, then under social distancing with parameter $\rho$ they would make $\text{Bin}(c_{\text{global}}, \rho)$ global contacts instead. We explore a uniform social distancing where the same contact reduction parameter is applied to every node, as an approximation to different levels of social distancing, in practice we expect there to be some dependence on household size. One person could do the shopping for several people, allowing for a greater reduction is global contacts compared to a one person household, or larger household may contain more key workers on average, reducing the households ability to socially distance. We observe this in the results of the CoMiX study [5], which we make use of when modelling different relaxations of lockdown measures. We can apply a social distancing measure dependent on the node's household size $\boldsymbol{\rho} = (\rho_1, \ldots, \rho_6)$ where a node's global contacts is given by $\text{Bin}(c_{\text{global}}, \rho_h)$. In doing so we can consider more realistic scenarios of social distancing, however in order for them to be meaningful we have to assume an underlying scenario, which we do arbitrarily.

| **Scenario** | **Workplace** | **School** | **Leisure** |
|---|---|---|---|
| A | 20% increase in contacts | 10% of contacts resume | 0% of contacts resume |
| B | 30% increase in contacts | 25% of contacts resume | 10% of contacts resume |
| C | 30% increase in contacts | 50% of contacts resume | 10% of contacts resume |
| D | 40% increase in contacts | 60% of contacts resume | 30% of contacts resume |
| E | 50% increase in contacts | 100% of contacts resume | 75% of contacts resume |

Table 3: Assumed scenarios of lockdown relaxation. The effect values are relative to the current lockdown number of contacts

We assigned contacts different labels, as follows:

1. `local` - frequency: "daily", setting: "at home", age: age of contact matches age of another household member
2. `global` - not a `local` contact
3. `work_travel` - a `global` contact with setting: "work" or "travel", age: participant age > 18
4. `school` - a `global` contact, age: participant age < 18, setting: "school"
5. `leisure` - a `global` contact, not `school`, not `work`, setting: "leisure"
6. `other` - a `global` contact, not `school` or `work` or `leisure`, setting: "otherplace"

We use data shared by the CoMiX study[5] to understand how global contacts have reduced in lockdown, however we are unable to share the data. We were able to estimate the reduction in each contact type due to the lockdown, and therefore in the case of school and leisure contacts we allowed them to resume.

### 4 Infection Model

The generation time is defined as the time from becoming infected to transmitting the infection. Getting an appropriately calibrated generation time distribution is important as the it defines the speed of the epidemic in relation to the speed of contact tracing. Our infection process is in terms of contacts, so contacts have a probability $\beta(\tau)$ of spreading the infection, where $\tau$ is the infectious age of a node, and we aim to set $\beta(.)$ such that it gives rise to a Weibull (mean=5, sd=1.92 days) distribution. We will consider $\beta_L(.)$ the local infection probabilities, and $\beta_G(.)$ the global infection probabilities, since we desire to calibrate an epidemic that gives the desired levels of local and global transmission for our model.

The generation times for transmissions that occur later in the local epidemic do not have the same generation distribution compared the those that occurred early in the local epidemic. The first local transmission only has a single infector, whereas the later transmission have multiple infectors, which biases the generation time towards smaller values. However, when generation times are estimated, they do not include infections that occur later in a household epidemic - the presence of multiple potential infectors makes it impossible to determine a generation time. As such, we calibrate $\beta_L(.)$ to give rise to correct pairwise generation time distribution, which is the distribution that will be estimated in practice.

Let $\tau$ be the infectious age of a node when the node infects a given local neighbour, and let $t^* = \tau|\{\tau < \infty\}$. Then $t^* \sim G^*$ where $G^*$ is drawn from the pairwise generation time distribution, and the rate of infection conditional upon $t_i < \infty$ is given by the hazard rate $h^*(t)$ of $G^*$. Define $f^*(t)$ and $S^*(t) = 1 - F^*(t)$ to be the conditional pdf and conditional survival functions of $G^*$, with $S(\infty) = \mathbb{P}(t_i = \infty)$. Then we have that

$$f^*(t) = \frac{f(t)}{1 - S(\infty)}, \quad S^*(t) = \frac{S(t) - S(\infty)}{1 - S(\infty)} \tag{3}$$

where $f(t)$ and $S(t)$ are the unconditional density and survival functions respectively. This leads to an expression for the unconditional hazard rate function in terms of the conditional pdf and survival function

$$h(t) = \frac{f(t)}{S(t)} = \frac{f^*(t)(1 - S(\infty))}{S^*(t)(1 - S(\infty)) + S(\infty)} \tag{4}$$

Therefore, if the pairwise hazard rate in our model equals $h(t)$ we obtain a rate of infection that gives rise to the pairwise generation time distribution. Increasing $S(\infty)$ increases the pairwise probability of an infection occurring and as such is one of the parameters we need to calibrate.

As our model progresses through discrete time steps, and the generation time distribution is continuous, we approximate the hazard rate as

$$\hat{h}(T) = \mathbb{P}(t_i = T | t_i > T - 1) = \int_{T-0.5}^{T+0.5} h(s) ds \tag{5}$$

Where $\hat{h}(T)$ is the discrete time hazard rate. In the event that $T = 0$, we take the lower limit of the integral to be 0.

That is any process which meets the above requirements gives rise to correctly distributed generation times. In our model, for an node with infectious age $t$ the probability of an infectious contact is $p\beta_L(t)$ where $p$ is the probability of a contact occurring and $\beta(t)$ is the probability that the a contact made by a node with infectious age $t$ will spread the infection. As such we aim to set $\beta(t)$ so that we obtain $\hat{h}(t)$.

Using standard discrete time survival analysis, we must satisfy

$$\hat{h}(T) = \frac{p\beta_L(T)}{\prod_{j=0}^{T-1}(1 - p\beta_L(j))}, \quad T > 0 \tag{6}$$

$$\hat{h}(T) = p\beta_L(0), \quad T = 0 \tag{7}$$

This system of equations can be solved recursively for $\beta_L(T)$, and for a given $S(\infty)$ and assumed distribution on $G^*$ with known hazard rate $\hat{h}(T) = \int_{T-0.5}^{T+0.5} h(s) ds$ can be rapidly evaluated.

In the global setting, we do not consider any depletion of a node's outside household contacts. Therefore in order to give rise to the correct generation time distribution for global contacts, let $t$ be the infectious age of the node making a global contact. Then

$$\beta_G(t) = \alpha f^*(t)$$

where $f^*$ is the probability density function of the pairwise generation time distribution. We note that $\alpha$ is a parameter to be tuned, as it controls the expected number of global infections.

This system of infection probabilities allows for well informed models of social distancing, and can even be used to handle increased within household contacts due to lockdown measures. We do not observe the frequency of local contacts increasing in the CoMiX data, but it is possible that the duration and closeness of contacts increases which may be more important for transmission. We used data from week 5 of CoMix.

### 5 Progression of the infection in a host

We model 3 epochs in the progression of the virus in a node's infection: time of infection to symptom onset $t_i$; from symptom onset to time of discovery $t_s$; and from time of discovery until recover, $t_d$. Time of infection $t_i$ records when a node became infected, the infectious age of a node is relative to this time.

If a node has been contact traced and has a symptom onset, some actions may be taken at symptom onset, depending on the model assumptions. For exposed nodes discovered through contact tracing and are therefore known to have been exposed to the virus, symptom onset allows further actions to take place, such as testing or propagation of contact tracing depending on the model assumptions.

Once the virus has been confirmed in a household through testing or symptom onset, the contact tracing process will propagate again, see section 6.3 for details.

Time of detection, $t_d$, follows symptom onset. In the case that a household is not under surveillance, this is when a node's symptoms are reported to a medical authority, which leads to; testing, isolation of the suspected case, quarantine of the other household members and contact tracing when the infection is confirmed. Not all infections are reported to a medical provider, due to either being asymptomatic or with mild symptoms that are disregarded, therefore we consider $\mathbb{P}(t_d < \infty) = \mathbb{P}(\text{infection discovery})$. The probability of infection discovery is difficult to estimate, and we vary across a prior distribution of beliefs.

The time delay from symptom onset to discovery is not known for the United Kingdom. The policy since March 12, 2020 for suspected Covid-19 infections has been to self isolate, but not necessarily report the infection and initiate contact tracing, which was stopped for most cases after this time. We were unable to reach a conclusion using FF100 data and as such we must turn to other sources to understand this delay. In [12] Pellis et al. estimate the UK symptom onset to hospitalisation to be mean 5.14 with standard deviation 4.20.

In Singapore however, Pellis et al. estimate the onset to visiting a medical provider was much shorter, with mean = 2.62 and standard deviation = 2.38. We expect this to be due to Singapore's past experience with SARS-CoV-1 and actively encouraging potential infections to be reported [12] and performing hospital quarantine of SARS-CoV-2 cases. We assume that this is the best representation of the delay from symptom onset to symptom reporting going forwards.

| **Parameter** | **Distribution** | **Source** |
|---|---|---|
| Delay from infection to symptom onset | Gamma($\mu$=4.83 days, $\sigma$=2.78) | [10] |
| Probability of infection discovery | 0.1, 0.2, 0.3, 0.4, 0.5 | Bounded using asymptomatic probabilities |
| Delay from symptom onset to infection discovery | Gamma($\mu = 2.62$, $\sigma = 2.38$) | [12] |

Table 4: Parameters and delays associated with the progression of the virus in a host.

#### 5.1 The importance of discovery

The probability of infection discovery is a parameter that is of crucial importance in our model, as it initiates household isolation and contact tracing events. When household quarantines are included in a model, the expected number of secondary infections caused by later generation of the household epidemic is reduced on average, as the infection may have been discovered in an earlier generation of the household epidemic, leading to quarantine of the household. As the likelihood of the virus being discovered increases, the number of household quarantines increases and the earlier in the local epidemic they occur, making it increasingly difficult for later generations of the virus to spread the infection. Additionally, if there is a contact tracing process this will lead to the process being initiated earlier in the epidemic, and more frequently. Household quarantine policies were announced in the UK on March 16th 2020, and as a result we believe the observed growth rate to have been impacted by this.

Later we discuss tuning the model to a growth rate, but the above discussion motivates the necessity of tuning the model under different assumptions of infection discovery, as infection discovery has a direct impact upon the growth rate of the epidemic.

The probability of infection discovery is a difficult one to estimate, consider asymptomatic and mild infections to be good indicators of the infection discovery probability. The definition of asymptomatic or mild infections change between studies, and an asymptomatic case can still be reported. In the town of Vo, Italy an extended effort was made to test the entire population [7], with 43.8% of cases being asymptomatic.

### 6 Contact Tracing Model

Contact tracing and quarantine is one of the most common interventions for epidemics. When an individual with an infection is discovered, attempts are made to trace their contacts who have been exposed and potentially infected. When a contact is successfully traced, they are quarantined to stop or reduce spread of the virus, which leads to a reduction in $R_{\text{eff}}$.

#### 6.1 Quarantine

A contact tracing process begins when an infection reports it's symptoms at time $t_d$, which immediately quarantines all nodes in the household. While a node is quarantined they are prevented from making global contacts, thus removing their ability to spread the infection globally, but local within household contacts continue unchanged and the local epidemic continues.

Let $Q$ be the day that a household enters quarantine and $S$ the day that a node in that household develops symptoms, then release from quarantine occurs according to the following rules [2];

$$\text{Time of release} = \begin{cases} Q + 14 & \text{if a node develops no symptoms} \\ S + 7 & \text{if a node develops symptoms on day S} \end{cases}$$

As such, it is possible for an infected node to be released from quarantine, if their symptoms onset after 14 days. We do not expect incubation periods of length >14 days, but it is possible for the node to have been infected shortly before their release, depending on the transmission chain in the household.

#### 6.2 Adherence to Quarantine

Evidence of uptake and adherence over time to individual and household quarantine orders from previous epidemics has been very mixed [16]. While the CoMiX survey has suggested that most of the population in the UK has followed lockdown policies, it is plausible that some individuals will be unable (e.g for economic reasons) or unwilling to follow quarantine notices that they receive. Particularly, as sections of society are reopened and contact tracing efforts intensify, there is a possibility that some individuals will be asked to repeatedly quarantine. Over time this may lead to non-adherence with quarantine measures which reduces the effectiveness of contact tracing.

We assume that all non-adherence is clustered by households, therefore households are assigned a propensity to non-adherence to quarantine measures with some probability. If the probability is one then this reduces to the case of nodes not adhering independent of any household structure. Nodes that do not belong to a household with the propensity to not adhere will always follow the rules, whereas node in households with the propensity to not adhere may choose to break the rules with some probability.

The simplest model of non-adherence is non-uptake of quarantine. Given that a node is in a household with the propensity to not adhere, then there is some probability that a node will continue making global contacts as normal when it should be quarantined.

The second model of non-adherence to quarantine is where nodes do uptake quarantine, however they leave early. Given that a node is in a household with the propensity to non-adherence, then each day of quarantine there is a probability $p_{leave}$ that the node will decide to leave early, leading to length of stay $\nu \sim \text{Geom}(p_{leave})$

$$\text{Time of release} = \begin{cases} \min(Q+14, Q+\nu) & \text{if a node develops no symptoms} \\ \min(S+7, Q+\nu) & \text{if a node develops symptoms on day S} \end{cases} \tag{8}$$

#### 6.3 Propagation of the Contact Tracing Process

The contact tracing process operates at the level of households, so we consider the household hypergraph induced by infection branching process on the individuals. Let $\mathcal{H}(t) = (V_H(t), E_H(t))$ where $N_H(t) = (h_1, \dots, h_n)$ the set of infected household nodes at time $t$ and $E^H(t) = [e^H_{ij}]$ the household edge matrix where

$$e^H_{ij} = \begin{cases} 1 & \text{if there is transmission between } i \text{ and } j \\ 0 & \text{otherwise} \end{cases} \tag{9}$$

As such if we allow backwards tracing, the edge matrix is undirected and the contact tracing can iterate backwards along the network, which can be potentially very useful. For example, it can allow for the discovery of sibling infections. If a node has many offspring, then there is an increased probability the node will be backwards traced by one of it's offspring. This means that contact tracing can then trace forwards again

A contact tracing process is initiated when an infection is discovered in a household which occurs according to the times of infection discovery of the nodes within the household. The household is immediately quarantined, reducing the ability of all nodes within the household to transmit globally. Next, the contact tracing process attempts to propagate along $\mathcal{H}(t)$. If $H$ is the household being quarantined at time $t$, then attempt to contact trace all neighbours of $H$, given by $\mathcal{N}(H)$. It is possible that some of these households will already be quarantined, in which case they are ignored. Suppose that $H$ attempts to trace $H' \in \mathcal{N}(H)$, the contact tracing attempt succeeds with $\mathbb{P}(\text{Contact tracing success}) = \gamma$, and if successful a contact tracing delay is drawn. Once the delay has passed, the nodes of $H'$ enter the quarantine state and surveillance state. Nodes in the surveillance state are aware that they, or someone else in the household, have been exposed to Covid-19 and as soon as there is symptom onset, the contact tracing process propagates again. It is possible for the contact tracing process to reach a household with symptom onset and immediately propagate again.

#### 6.4 Delays, success probabilities and strategy in contact tracing

It is not always possible for a contact tracing attempt to succeed for a multitude of different reasons. Not all contacts are recalled, some are strangers who cannot be identified and it may be impossible to reach others. Additionally, contact tracing is not an instant process and the speed at which it progresses is determined by the nature of the infection and by the delays in contact tracing attempts between households.

The true success probability of a contact tracing process cannot be easily known. Even if you are able to trace 100% of contacts reported by an infection, these are only the contacts that they report, so the true number of contacts that they made may be higher. In the UK, approximately 90% of contact tracing attempts were

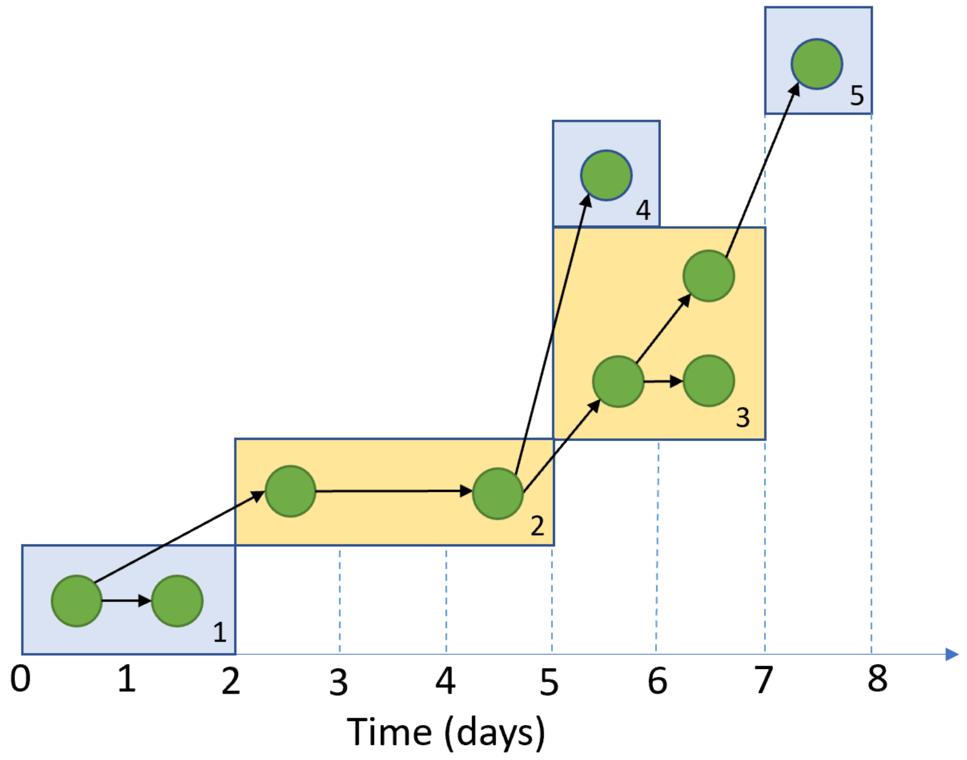


Figure 1: A demonstration of a household branching process with a contact tracing process, the households are identified by numbers in the bottom right hand corner of each rectangle. The infection is discovered in case 4. This quarantines household 2 and initiates contact tracing of connected households. The backwards tracing attempt to household 1 succeeds, with a time delay of 2 days. Household 3 is traced immediately, quarantining several cases early in their infection. When there is symptom onset in one of these cases the contact tracing process will propagate again by attempting to reach household 5, potentially after a testing delay. Household 4 was not successfully traced by household 2, and will continue to spread the infection. The x-axis refers to the temporal evolution of the transmission process in this example.

| Parameter | Distribution | Source |
|---|---|---|
| Tracing delay distribution | Poisson($\lambda_{delay}$) | Assumed |
| Tracing delay parameter | $\lambda_{delay} \sim$ Uniform(1.5, 2.5) | Estimates from PHE anonymised line-list data |
| Tracing success probability | Uniform(0.7, 0.95) | [1] |

Table 5: Parameters associated with the contact tracing process

successful, so we have chosen to vary the contact tracing success probability from 70% to 90%. We have assumed that tracing delays are distributed Poisson, with the mean tracing delay varying from between 1.5 to 2.5 days.

As for the tracing delay, we believe the tracing delay to be Poisson distributed with a mean delay of 2 days. We have chosen to vary this mean delay between 1.5 and 2.5.

Contact tracing is only propagated when an infection is discovered, or when symptoms onset in a house that is under symptom surveillance. This sets a form of speed limit for a contact tracing process, and one that can be particularly problematic for SARS-CoV-2, given that many infections occur before symptom onset. It is possible that a positive test may be confirmed before the contact tracing process is able to propagate, which adds further delays to the process.

One approach to overcoming the speed limit in contact tracing is two-step tracing. The index of a household is defined to be the distance it is away from a confirmed infection, so households with a confirmed infection are index 0. The idea with two step tracing, is to immediately propagate contact tracing when the process reaches an index 1 household (i.e; do not wait for symptom onset), because nodes in the index 1 household may be infected and may have already spread the infection.

##### 6.4.1 Digital Contact Tracing

A wide variety of contact tracing applications are in development which aim to improve contact tracing by increasing the probability of a successful contact tracing, and/or the speed at which the contact is traced. There are numerous difficulties in evaluating how effective a contact tracing app will be, and whether Bluetooth or location data is a good proxy for a contact in epidemiological terms.

We assume a 'perfect world' contact tracing application. If both ends of an edge have the contact tracing application, then the edge will be digitally contact traced, which guarantees contact tracing success and performs the contact tracing instantly. Additionally we assume that nodes have the app independent of any other factors. Therefore, the probability of two randomly picked individuals both having the contact tracing app, is quadratic in the probability of someone having the app. As we require both individuals on either end of tracing event to have the app for the tracing to be performed digitally, then the probability of a tracing being performed digitally quadratic in the probability that a randomly chosen individual has the app.

The assumption that nodes have the app independent of anything else is likely incorrect, a plausible alternative is that the distribution of the app across the population is such that a small number of households have high app uptake, which leads to an even lower probability of an edge being digitally contact traced.

### 7 Model Calibration and Results analysis

#### 7.1 Estimating the growth rate of a simulated epidemic

To simulate the growth rate of an epidemic we begin with 5000 infections on day 0. The choice of high starting infections is so that we can estimate exponential decay, as well as exponential growth, without prior knowledge beforehand. The simulation is then run for 20 days. Initially most epidemics experience rapid growth as they spread through their starting households with relative ease, slowing as susceptible depletion kicks in and as the contact tracing process is initiated by infection discovery. In practice we find that after

10 days the system enters a state of stable decay or growth and we therefore use days 10 to 20 to estimate the growth rate of the epidemic.

For a given epidemic let $y_t$ be the number of new transmissions that occur on day $t$. We fit a robust linear regression model of $\log(y_t) = \beta_0 + \beta_1 t$ where $\beta_1 = r$, the Malthusian growth rate of the epidemic and $y_0 = \exp(\beta_0)$ the initial rate of infection at $t = 10$. The robust linear regression to be necessary for the epidemics with large negative growth rates as $Var(y_t)$ increases as $y_t \to 0$.

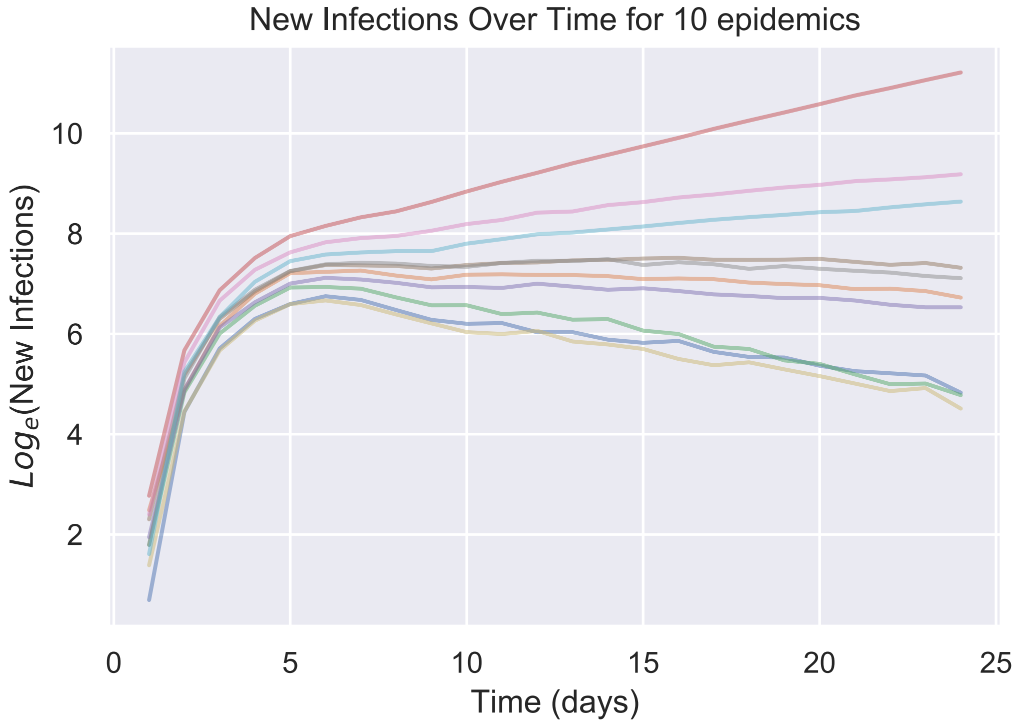


Figure 2: A demonstration of the initial rapid epidemic growth settling down to a constant growth rate after approximately 10 days, or roughly 3 generation times.

#### 7.2 Household Secondary Attack Rate

We aim to tune the parameters such that the model has a realistic household secondary attack rate, defined to be the proportion of susceptible household members not including the index case that become infected in a local epidemic. We review the results of different household studies in table 6 and we assume a household secondary attack rate of 21.4%, the mean of 3 studies that were available at the time.

As such we need to set $S_{local}(\infty)$, the pairwise survive forever probability of the local epidemic. It is computationally cheap to simulate the local epidemics, so we simulate 1000 epidemics for a range of proposal values, estimate the household secondary attack rate, and perform linear regression to obtain the value of $S_{\text{local}}(\infty)$. The household sizes are drawn from the size-mean-contacts biased distribution of households 2, which is the distribution of household sizes given that the household is infected and the distribution of household sizes we expect to see in practice.

| Estimated Household SAR | Source |
|---|---|
| 16.3% | [14] |
| 19.3% of those living at the same address | [6] |
| 30% | [15] |

Table 6: Estimates of the Household Secondary attack rate from the literature

##### 7.3 Epidemic Growth Rate

We desire to tune our model so that the baseline scenario has a growth rate of $r = 0.22$ [12], corresponding to a growth rate of just over 3 days and defining the baseline scenario to be an epidemic growing with, household quarantine upon infection, no contact tracing and no social distancing.

The growth rate of our model is determined by the triple

$$\theta = (S_{local}(\infty), \alpha, \mathbb{P}(\text{infection discovery}))$$

where $S_{local}(\infty)$ determines the pairwise survival probability within households, $\alpha$ scales the infectiousness of outside household contacts. Finally $\mathbb{P}(\text{infection discovery})$ is the probability of an individual discovering they are infected and reporting it, leading to household isolation. As discussed, this impacts upon epidemic dynamics since later generation of the household epidemic may be quarantine early in their infection due to infection discovery in another household member.

#### References


[1] PHE: COVID-19: Draft contact tracing operating model, 23 April 2020.

[2] Stay at home: guidance for households with possible coronavirus (COVID-19) infection. Library Catalog: www.gov.uk.

[3] Akira Endo, Mitsuo Uchida, Adam J. Kucharski, and Sebastian Funk. Fine-scale family structure shapes influenza transmission risk in households: Insights from primary schools in Matsumoto city, 2014/15. *PLOS Computational Biology*, 15(12):e1007589, December 2019. Publisher: Public Library of Science.

[4] Joel Hellewell, Sam Abbott, Amy Gimma, Nikos I. Bosse, Christopher I. Jarvis, Timothy W. Russell, James D. Munday, Adam J. Kucharski, W. John Edmunds, Fiona Sun, Stefan Flasche, Billy J. Quilty, Nicholas Davies, Yang Liu, Samuel Clifford, Petra Klepac, Mark Jit, Charlie Diamond, Hamish Gibbs, Kevin van Zandvoort, Sebastian Funk, and Rosalind M. Eggo. Feasibility of controlling COVID-19 outbreaks by isolation of cases and contacts. *The Lancet Global Health*, 8(4):e488–e496, April 2020. Publisher: Elsevier.

[5] Christopher I. Jarvis, Kevin Van Zandvoort, Amy Gimma, Kiesha Prem, Petra Klepac, G. James Rubin, and W. John Edmunds. Quantifying the impact of physical distance measures on the transmission of COVID-19 in the UK. *BMC Medicine*, 18(1):1–10, December 2020. Number: 1 Publisher: BioMed Central.

[6] Qin-Long Jing, Ming-Jin Liu, Jun Yuan, Zhou-Bin Zhang, An-Ran Zhang, Natalie E. Dean, Lei Luo, Meng-Meng Ma, Ira Longini, Eben Kenah, Ying Lu, Yu Ma, Neda Jalali, Li-Qun Fang, Zhi-Cong Yang, and Yang Yang. Household Secondary Attack Rate of COVID-19 and Associated Determinants. *medRxiv*, page 2020.04.11.20056010, April 2020. Publisher: Cold Spring Harbor Laboratory Press.

[7] Enrico Lavezzo, Elisa Franchin, Constanze Ciavarella, Gina Cuomo-Dannenburg, Luisa Barzon, Claudia Del Vecchio, Lucia Rossi, Riccardo Manganelli, Arianna Loregian, Nicolò Navarin, Davide Abate, Manuela Sciro, Stefano Merigliano, Ettore Decanale, Maria Cristina Vanuzzo, Francesca Saluzzo, Francesco Onelia, Monia Pacenti, Saverio Parisi, Giovanni Carretta, Daniele Donato, Luciano Flor,

Silvia Cocchio, Giulia Masi, Alessandro Sperduti, Lorenzo Cattarino, Renato Salvador, Katy A. M. Gaythorpe, Imperial College London COVID-19 Response Team, Alessandra R. Brazzale, Stefano Toppo, Marta Trevisan, Vincenzo Baldo, Christl A. Donnelly, Neil M. Ferguson, Ilaria Dorigatti, and Andrea Crisanti. Suppression of COVID-19 outbreak in the municipality of Vo, Italy. *medRxiv*, page 2020.04.17.20053157, April 2020. Publisher: Cold Spring Harbor Laboratory Press.

[8] Zachary J. Madewell, Yang Yang, Ira M. Longini, M. Elizabeth Halloran, and Natalie E. Dean. Household transmission of SARS-CoV-2: a systematic review and meta-analysis of secondary attack rate. preprint, Infectious Diseases (except HIV/AIDS), August 2020.

[9] Joël Mossong, Niel Hens, Mark Jit, Philippe Beutels, Kari Auranen, Rafael Mikolajczyk, Marco Massari, Stefania Salmaso, Gianpaolo Scalia Tomba, Jacco Wallinga, Janneke Heijne, Malgorzata Sadkowska-Todys, Magdalena Rosinska, and W. John Edmunds. Social Contacts and Mixing Patterns Relevant to the Spread of Infectious Diseases. *PLoS Medicine*, 5(3):e74, March 2008.

[10] Christopher E. Overton, Helena B. Stage, Shazaad Ahmad, Jacob Curran-Sebastian, Paul Dark, Rajenki Das, Elizabeth Fearon, Timothy Felton, Martyn Fyles, Nick Gent, Ian Hall, Thomas House, Hugo Lewkowicz, Xiaoxi Pang, Lorenzo Pellis, Robert Sawko, Andrew Ustianowski, Bindu Vekaria, and Luke Webb. Using statistics and mathematical modelling to understand infectious disease outbreaks: COVID-19 as an example. *arXiv:2005.04937 [physics, q-bio]*, May 2020. arXiv: 2005.04937.

[11] Lorenzo Pellis, Frank Ball, and Pieter Trapman. Reproduction numbers for epidemic models with households and other social structures. I. Definition and calculation of R0. *Mathematical Biosciences*, 235(1):85–97, January 2012.

[12] Lorenzo Pellis, Francesca Scarabel, Helena B. Stage, Christopher E. Overton, Lauren H. K. Chappell, Katrina A. Lythgoe, Elizabeth Fearon, Emma Bennett, Jacob Curran-Sebastian, Rajenki Das, Martyn Fyles, Hugo Lewkowicz, Xiaoxi Pang, Bindu Vekaria, Luke Webb, Thomas A. House, and Ian Hall. Challenges in control of Covid-19: short doubling time and long delay to effect of interventions. *medRxiv*, page 2020.04.12.20059972, April 2020. Publisher: Cold Spring Harbor Laboratory Press.

[13] Gail E. Potter, Mark S. Handcock, Ira M. Longini, and M. Elizabeth Halloran. ESTIMATING WITHIN-HOUSEHOLD CONTACT NETWORKS FROM EGOCENTRIC DATA. *The annals of applied statistics*, 5(3):1816–1838, 2011.

[14] Li W, Zhang B, Lu J, Liu S, Chang Z, Cao P, Liu X, Zhang P, Ling Y, Tao K, and Chen J. The Characteristics of Household Transmission of COVID-19, April 2020. ISSN: 1537-6591 Library Catalog: pubmed.ncbi.nlm.nih.gov Publisher: Clin Infect Dis.

[15] Zhongliang Wang, Wanli Ma, Xin Zheng, Gang Wu, and Ruiguang Zhang. Household transmission of SARS-CoV-2. *Journal of Infection*, April 2020.

[16] Rebecca Webster, Samantha Brooks, Louise Smith, Lisa Woodland, Simon Wessely, and James Rubin. How to improve adherence with quarantine: Rapid review of the evidence. *medRxiv*, page 2020.03.17.20037408, March 2020. Publisher: Cold Spring Harbor Laboratory Press.
